# Developmental origins of exceptional health and survival: A four-generation family cohort study

**DOI:** 10.1101/2024.05.04.24306872

**Authors:** Matthew Thomas Keys, Dorthe Almind Pedersen, Pernille Stemann Larsen, Alexander Kulminski, Mary F. Feitosa, Mary Wojczynski, Michael Province, Kaare Christensen

## Abstract

Descendants of longevity-enriched sibships demonstrate a broad health and survival advantage throughout the life course. However, little is known about manifestations during very early life. Here we show a pattern of lower risk of adverse early life outcomes in third-generation grandchildren (N = 5637) of Danish longevity-enriched sibships compared to the general population, including infant mortality (Hazard Ratio = 0.53, 95% CI [0.36, 0.77]) and a range of neonatal health indicators. These associations in fourth-generation great-grandchildren (N = 14,908) were strongly attenuated and less consistent (e.g., infant mortality, Hazard Ratio = 0.90, [0.70, 1.17]). These dilatory patterns across successive generations were independent of stable socioeconomic and behavioural advantages (e.g., parental education and maternal smoking), maternal and paternal lines of transmission, as well as secular trends in the background population. Our findings suggest that exceptional health and survival may have early life developmental components and implicate heritable genetic and or epigenetic factors in their transmission.

**Background:** Previous researched has demonstrated potent health and survival advantages across three-generations in longevity-enriched families. However, the survival advantage associated with familial longevity may manifest earlier in life than previously thought.

**Methods:** We conducted a matched cohort study comparing early health trajectories in third-generation grandchildren (n = 5,637) and fourth-generation great-grandchildren (n = 14,908) of longevity-enriched sibships to demographically matched births (n = 41,090) in Denmark between 1973 and 2018.

**Results:** Lower risk was observed across a range of adverse early life outcomes in the grandchildren, including infant mortality (Hazard Ratio (HR) = 0.53, 95% CI [0.36, 0.77]), preterm birth (Odds Ratio (OR) = 0.82, [0.72, 0.93]), small for gestational age (OR = 0.83, [0.76, 0.90]) and neonatal respiratory disorders (OR = 0.77, [0.67, 0.88]). Relative advantages in parental education and maternal smoking were observed in both generations to a similar degree. However, a much smaller reduction in infant mortality was observed in the great-grandchildren (HR = 0.90, [0.70, 1.17]) and benefits across other outcomes were also less consistent, despite persisting socioeconomic and behavioural advantages. Lastly, maternal, and paternal lines of transmission were equipotent in the transmission of infant survival advantages.

**Conclusions:** Descendants of longevity-enriched sibships exhibit a broad health advantage manifesting as early the perinatal period. However, this effect is strongly diluted over successive generations. Our findings suggest that exceptional health and survival may have early developmental components and implicate heritable genetic and or epigenetic factors in their specific transmission.

**Key Messages:**

- Previous researched has demonstrated potent health and survival advantages across three-generations in longevity-enriched families. However, the survival advantage associated with familial longevity may manifest earlier in life than previously thought.
- In our study of third and fourth-generation descendants of longevity-enriched sibships, we observed a broad infant health and survival advantage reflected by protection against a diverse range of adverse birth outcomes.
- These advantages were strongly attenuated between the third and fourth generations, independent of otherwise stable socioeconomic and behavioural parental advantages, as well as maternal and paternal lines of transmission.
- Our findings suggest that familial aggregation of exceptional health and survival may have early life developmental components and triangulate to implicate heritable genetic and or epigenetic factors in their transmission.

## Introduction

The study of long-lived persons is of key importance for understanding the mechanisms that confer protection against age-related morbidity.^1^ Replicating their example more broadly in the face of aging populations has the potential to bring powerful social and economic benefits.^2,3^ However, the diversity of aging trajectories within and across populations reflects the broad spectrum of its environmental and genetic determinants.^4^ Studies assessing the combined impact of behavioral risk factors in mid- and late-life estimate that these alone may account for up to 10 to 14 years of life expectancy.^5–8^ Studies of twins suggest a relatively modest contribution of genetic factors to variation in overall lifespan.^9^ However, these factors likely become increasingly important in survival at advanced ages, which is further evidenced by the familial aggregation of exceptional longevity.^10,11^ Studies of families selected for their aggregation of longevity are expected to play a key role in advancing our understanding of genetic and environmental determinants of exceptional health and survival.^12–14^

One area that has received less attention is the impact of life-course history prior to mid-life, and especially the early developmental periods.^15^ Life-course studies in the context of exceptional longevity are often impractical due to length of follow-up, historical data and contemporary comparison cohorts required.^15^ However, descendants of long-lived persons and longevity-enriched sibships often display similar health and metabolic profiles to their ancestors and can serve as a model for healthy ageing.^16,17^ Multigenerational family studies in particular have proved useful for providing opportunities to circumvent these methodological challenges.^18^ Previous findings by our group have demonstrated potent health and survival advantages across three-generations of longevity-enriched families and implicated behavioural mechanisms in the familial aggregation of this phenotype.^19–22^ We also observed indications that the survival advantage associated with familial longevity may manifest earlier in life than previously thought, suggesting a possible developmental component to exceptional longevity.^21^ However, the robustness, mechanisms and implications of this finding, broader health manifestations in early life, as well as transmission to subsequent generations, are currently unclear.

In this study we utilised a multigenerational cohort of longevity-enriched sibships and their descendants in Denmark to broadly assess early developmental health trajectories associated with the familial aggregation of longevity. Administrative registers covering all residents of Denmark facilitated linkage of our cohort to its fourth- generation descendants and comparisons with the general population. Clinical registries established in the 1970s permitted measurement of a range of neonatal, infant, and maternal health phenotypes in third generation (G3) grandchildren and fourth generation (G4) great-grandchildren of our cohort. We compared these to demographically matched births from the general population, assessed the transmission of developmental effects over multiple generations, and explored mediating roles of maternal and paternal, socioeconomic, and behavioural factors. Lastly, we interpreted our findings in the context of research examining the relationship between early life health trajectories and adult and late-life health phenotypes.

## Materials and Methods

### Danish Longevity-Enriched Families

In this study we utilised a Danish cohort of 659 families selected for their aggregation of exceptional longevity.^21^ The cohort is comprised of three consecutive studies, including the Danish Oldest Siblings pilot study, the Genetics of Healthy Aging study, and the Danish participants of the National Institute of Aging’s Long Life Family Study (LLFS).^13^ Overall, the cohort’s families contained two or more siblings that reached 88 ≥ years of age and were alive at the time of recruitment between 2006-2009. However, 99.5% had two or more siblings in the proband generation who survived until at least 90 years of age.

In collaboration with the LLFS, this larger cohort of 659 Danish families (78 of whom participate directly in LLFS) has been employed as an ancillary resource to further study exceptional aging. The cohort facilitates population-based follow-up of longevity-enriched families with minimal attrition as well as comparisons with the general population through a system of national registries. Ascertainment of 2^nd^ generation (G2) offspring and 3^rd^ (G3) generation grandchildren through the Danish Civil Registration System has been described elsewhere and feature in a range of published research activities.^21,23^ In this study we extended the cohort through the same process to include the 4^th^ generation (G4) great-grandchildren (See Figure 1).

**Figure 1.**
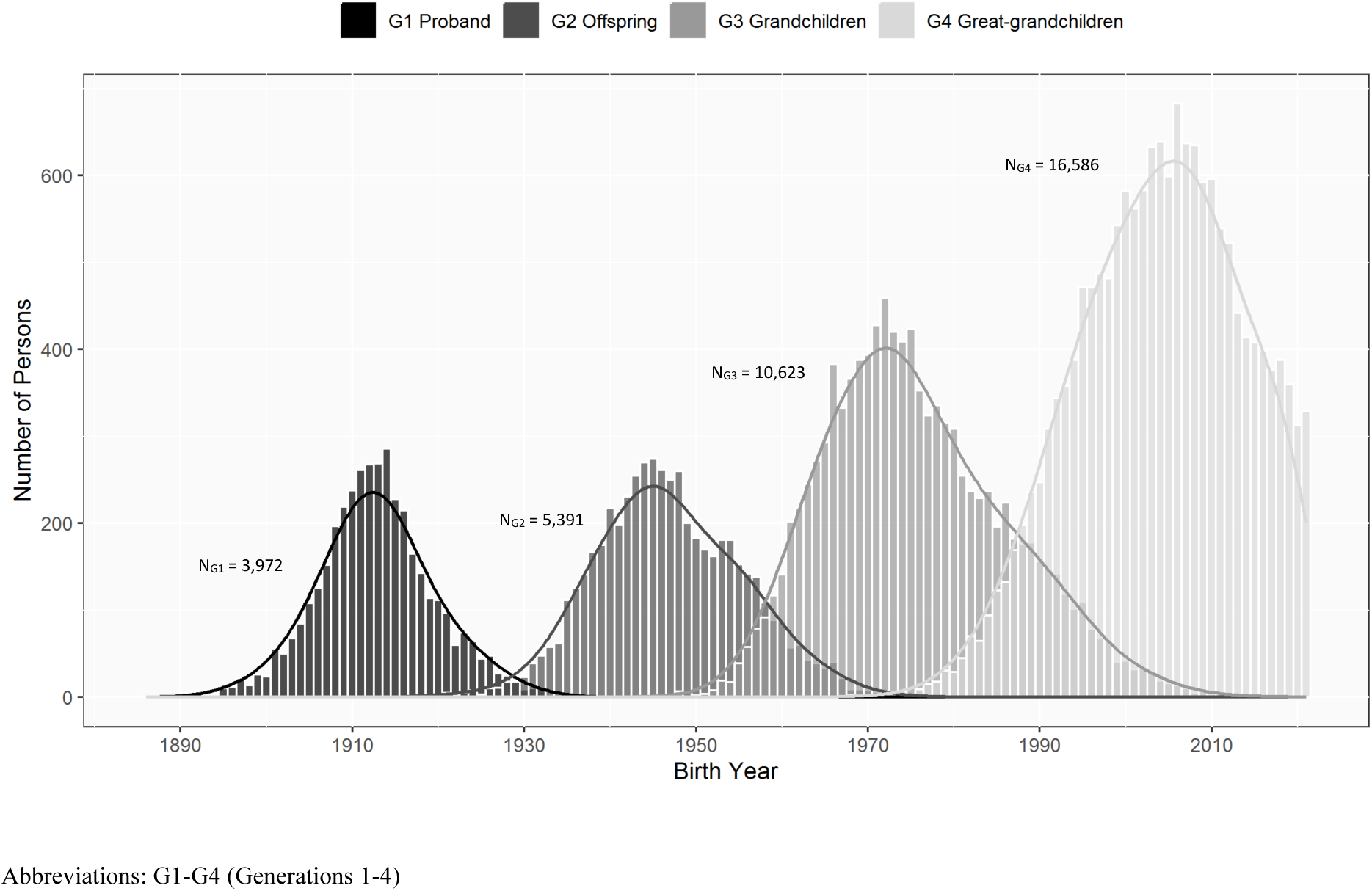
Distribution by birth year of successive generations descending from a proband cohort of longevity-enriched sibships

### Study Design

We performed a matched cohort study in the Danish population between 1^st^ January 1973 to 31^st^ December 2018. The Danish Medical Birth Registry was used to identify and extract information on live births in Denmark since 1973.^24^ The Danish National Patient Registry was used to ascertain additional conditions associated with childbirth, based on diagnoses listed on inpatient hospitalisations since 1977.^25^ The Civil Registration System (CRS) was used to determine status across follow-up, including death and emigration.^23^ Data on parental education history was obtained from the Population Education Register, with coverage exceeding 90% of the population.^26^ Education levels were defined according to the 2011 International Standard Classification of Education and coded as the highest attained level between the mother or father in our analyses.

Our source population included all live births in Denmark between 1^st^ January 1973 and 31^st^ December 2018 with complete data on parity, maternal birth date, and maternal country of birth. Live births from Danish LEFs were compared to demographically matched live births from the general population at a 1:2 ratio. Matching criteria included sex, birth year, birth season, maternal birth year, maternal parity and whether the mother was born in Denmark or abroad. Participants who couldn’t be matched on these criteria were removed (see Sensitivity Analyses).

### Outcomes

Our primary outcome was infant mortality, defined as death within the first 365 days of life observed in the Civil Registration System. Secondary neonatal outcomes included preterm birth, small for gestational age, large for gestational age, low Apgar score, birth trauma, neonatal respiratory disorders, congenital malformations, and other neonatal morbidity. We also assessed several maternal outcomes including assisted delivery, caesarean section delivery, preeclampsia and eclampsia, placental disorders, haemorrhage, and other maternal morbidity. Outcomes for ‘other neonatal morbidity’ and ‘other maternal morbidity’ were included in our study to provide the broadest scope for assessing health differences and included all adverse conditions related to the pregnancy, delivery and health of the neonate outside of those listed above (see Supplementary Material Table S1.2).

Preterm birth was defined as delivery prior to the end of 37^th^ week of gestation. Small and large for gestational age were defined as belonging to the below the 10^th^ and above the 90^th^ percentiles respectively of birth weights for gestational age calculated from intrauterine growth curves based on ultrasonically estimated foetal weights.^27^ Low Apgar score was defined as a value of below 7 as measured after 5 minutes following delivery. Outcomes based on gestational age, weight for gestational age, and Apgar scores were available since the initiation of the Medical Birth Registry in 1973.^24^ All remaining neonatal and maternal outcomes were based on inpatient diagnoses observed in the National Patient Registry and were only available since 1977. Neonatal events were defined as those occurring in the first 28 days of life, and maternal events as occurring at any point during pregnancy, delivery, or the puerperium (up to 6 weeks following delivery). Full ICD-8 (1973-1992) and ICD-10 (1993-2018) codes used to define our outcomes and further explanations are presented in the Supplementary Material (Table S1.2).

We also assessed differences in maternal smoking and parental education levels between longevity-enriched families and the general population, as indicators of behavioural and socioeconomic characteristics. Maternal smoking during pregnancy is likely a causal determinant of a range of adverse neonatal and infant health outcomes, but may also serve as a proxy for other behavioural manifestations associated with poor health.^28,29^ Data on maternal smoking was only available since 1991 in the Medical Birth Registry and was classified as a binary variable indicating any amount of smoking in any of the trimesters of pregnancy, regardless of later cessation. Parental education data was obtained from the Population Education Register and coded with the following categories: ‘Primary or Lower Secondary’, ‘Upper Secondary’, ‘Short Cycle Tertiary’, ‘Bachelor or Equivalent’, ‘Master, Doctoral or Equivalent’.

### Statistical Analyses

Differences in outcomes between G3 grandchildren and G4 great-grandchildren of longevity-enriched sibships and matched controls were assessed by Cox proportional hazard, conditional logistic, multinomial and ordinal logistic regression models.

For analyses of infant mortality, matched sets were followed up until whichever came first out of death, emigration, 365 days after birth, or the end of the observed data and analysed by Cox regression models. For analyses of neonatal and maternal morbidities, as well as maternal smoking, outcomes were treated as binary and estimated by conditional logistic regression models. For analyses of parental education level, the outcome was treated as categorical and were estimated by multinomial and ordinal logistic regression models.

All models accounted for the matching variables via stratification, and estimated robust standard errors clustered on maternal ID to account for several births involving the same mother. Where applicable, we also calculated separate estimates with further adjustment for parental education level at time of birth, except for analyses where this was the outcome. Analyses of neonatal and maternal morbidities as secondary outcomes were adjusted for multiple testing via the Hommel method, which assumes that outcomes are non-negatively correlated (n_outcomes_ = 14).^30,31^

Differences in maternal or paternal lines of transmission of infant phenotypes were assessed via interaction models. Separate estimates were provided for subgroups where the mother or father was a descendant of a longevity-enriched sibship compared to a marry-in. In the presence of no interactions, this would suggest an equal role for maternal and paternal factors in the transmission of exceptional survival and health in the neonatal and infant periods. We apply this methodology to analyses of infant survival and maternal smoking, and also to analyses of overall neonatal and maternal morbidities in the Supplementary Material (see Sensitivity Analyses).

Patterns of transmission of health and survival advantages across generations in longevity-enriched families were assessed by directly comparing G3 grandchildren and G4 grandchildren in calendar periods where these successive generations have overlapping birth cohorts (approx. 1973 – 2010). We performed regression analyses with statistical adjustment for the same covariates used as matching criteria when selecting general population controls. Since this included birth year, these analyses were robust to changes in secular trends in the background population over the same period. A range of supplementary analyses were conducted to assess the suitability and robustness of this approach (see Sensitivity Analyses).

All statistical analyses were conducted in the R Statistical Software (v4.2.3; R Core Team 2023). All confidence intervals and two-sided hypothesis tests were provided at 95% and 5% levels respectively.

### Sensitivity Analyses

We performed several sensitivity analyses to assess the robustness of our results. First, we assessed the impact of including children who were unable to be matched initially, by weaking their matching criteria and then appending them to our fully matched data and repeating key analyses. We then assessed longer windows of opportunity for the diagnosis of congenital malformations. We also examined mean differences in continuous rather than dichotomized outcomes where possible. Outcomes with incomplete data were also analysed to determine if their observations were missing at random and, if so, the direction of potential biases. We assessed robustness to adjustment for paternal country of birth and age at conception. For outcomes based on birth weight, we also evaluated more granular subgroups, for example very and extremely small for gestational age, and low birth weight in general. To capture broad patterns in neonatal and maternal morbidities, we assessed health advantages based on various composite scores comprised of the individual neonatal and maternal endpoints included in our study.

Lastly, we assessed the robustness of our ‘direct comparison’ methodology via use of several negative control analyses. These analyses tested whether our results were biased by the known secular trends of improved neonatal outcomes and infant survival throughout the study period. First, as described in the statistical analyses section, we directly compared G3 grandchildren to G4 great-grandchildren in calendar periods where these successive generations have overlapping birth cohorts (approx. 1973 – 2010). These analyses statistically adjusted for the same criteria used in our previous matched analyses, including birth year, birth season, sex, maternal age and parity. We then compared G3 matched controls to G4 matched controls using the same models where, if they adequately adjust for the confounding induced by matching (e.g. different distributions of maternal age and parity), no differences in survival should be observed. Several other comparisons were also conducted for completeness. More information on all these analyses can be found in Section 3 of the Supplementary Material.

## Results

### Study Population

Our source population included the entire population living in Denmark since 1968. After extending our cohort of longevity-enriched families to the great-grandchildren, we identified a total of 10,623 third-generation (G3) grandchildren (born between 1950 and 2010) and 16,586 fourth-generation (G4) great-grandchildren (born between 1970 and 2018) (See Figure 1). From this cohort, we selected live births occurring in Denmark between 1^st^ January 1973 and 31^st^ December 2018 and removed observations with invalid CPR numbers or incomplete data on matching criteria in the Medical Birth Registry. The resulting sample included 5,718 grandchildren and 14,968 great-grandchildren from longevity-enriched families (LEFs), and 980,232 potential control births from the general population.

After matching children from LEFs to controls from the general population at a 1:2 ratio, 141 were unable to be matched (see Sensitivity Analyses). Our final study population included a total of 61,635 live births from a total of 51,234 unique mothers. This included 5,637 grandchildren and 14,908 and great-grandchildren of longevity-enriched families and 41,090 matched controls. Figure S1.1 of the Supplementary Material describes the selection process used in this study in detail and loss of participants for each criterion.

Table 1 describes the baseline birth characteristics of the final matched cohorts analysed for this study. Minimal differences in maternal age at conception were due to matching on birth year. Parity was higher in G3 grandchildren (0.93) compared to G4 great-grandchildren (0.69) as we only included births since 1973, once the Danish Medical Birth Registry had been established.^24^ Births before this year were excluded and thus the G3 grandchildren included in our study were less likely to first child compared to the G4 great-grandchildren.

**Table 1.** Baseline characteristics of liveborn children and matched controls, stratified by grandchildren and great-grandchildren of longevity-enriched sibships.

| Characteristic | G3 Grandchildren |  | G4 Great-grandchildren |  |
| --- | --- | --- | --- | --- |
|  | LEF | Control | LEF | Control |
| Number of live births | 5637 | 11274 | 14908 | 29816 |
| Birth Year (n, %) |  |  |  |  |
| 1973-1976 | 1509 (26.8) | 3018 (26.8) | 22 (0.1) | 44 (0.1) |
| 1977-1980 | 1195 (21.2) | 2390 (21.2) | 87 (0.6) | 174 (0.6) |
| 1981-1984 | 890 (15.8) | 1780 (15.8) | 266 (1.8) | 532 (1.8) |
| 1985-1988 | 731 (13.0) | 1462 (13.0) | 594 (4.0) | 1188 (4.0) |
| 1989-1992 | 583 (10.3) | 1166 (10.3) | 1088 (7.3) | 2176 (7.3) |
| 1993-1996 | 378 (6.7) | 756 (6.7) | 1624 (10.9) | 3248 (10.9) |
| 1997-2000 | 194 (3.4) | 388 (3.4) | 1991 (13.4) | 3982 (13.4) |
| 2001-2004 | 98 (1.7) | 196 (1.7) | 2290 (15.4) | 4580 (15.4) |
| 2005-2008 | 39 (0.7) | 78 (0.7) | 2451 (16.4) | 4902 (16.4) |
| 2009-2012 | 13 (0.2) | 26 (0.2) | 2172 (14.6) | 4344 (14.6) |
| 2013-2018 | 7 (0.1) | 14 (0.1) | 2323 (15.6) | 4646 (15.6) |
| Birth Season (n, %) |  |  |  |  |
| Autumn | 1302 (23.1) | 2604 (23.1) | 3681 (24.7) | 7362 (24.7) |
| Spring | 1566 (27.8) | 3132 (27.8) | 3785 (25.4) | 7570 (25.4) |
| Summer | 1471 (26.1) | 2942 (26.1) | 3985 (26.7) | 7970 (26.7) |
| Winter | 1298 (23.0) | 2596 (23.0) | 3457 (23.2) | 6914 (23.2) |
| Maternal Age (sd) | 29.44 (4.71) | 29.42 (4.71) | 30.15 (4.53) | 30.15 (4.53) |
| Parity (sd) | 0.93 (0.90) | 0.93 (0.90) | 0.69 (0.79) | 0.69 (0.79) |
| Danish Mother (n, %) | 5514 (97.8) | 11028 (97.8) | 14376 (96.4) | 28752 (96.4) |
| Male (n, %) | 2824 (50.1) | 5648 (50.1) | 7661 (51.4) | 15322 (51.4) |
| Highest Parental Education (n, %) |  |  |  |  |
| Primary or Lower Secondary | 674 (12.1) | 1703 (15.2) | 767 (5.1) | 2437 (8.2) |
| Upper Secondary | 2476 (44.3) | 5272 (47.2) | 5629 (37.8) | 12282 (41.3) |
| Short cycle tertiary | 343 (6.1) | 548 (4.9) | 943 (6.3) | 2065 (6.9) |
| Bachelor or equivalent | 1458 (26.1) | 2601 (23.3) | 4307 (28.9) | 7780 (26.1) |
| Master or equivalent | 623 (11.1) | 1016 (9.1) | 3002 (20.1) | 4845 (16.3) |
| Doctoral or equivalent | 16 (0.3) | 25 (0.2) | 258 (1.7) | 363 (1.2) |
| Maternal Smoking Status |  |  |  |  |
| Current smoker | 192 (21.8) | 452 (26.0) | 1597 (12.9) | 4240 (17.1) |
| Not a current smoker | 689 (78.2) | 1285 (74.0) | 10803 (87.1) | 20543 (82.9) |
| LEF Mother (n, %) | 2432 (43.1) | - | 7786 (52.2) | - |

238 Abbreviations: G3 (Generation 3), G4 (Generation 4), LEF (Longevity-enriched Family)

Figure 1 describes the distribution of birth years in all four generations of LEFs in our source cohort. Years 1973- 2010 represent a period of overlap between the two generations also with observation time in our study. We use this overlapping period to assess the transmission of survival and health patterns across generations in longevity-enriched families, and perform analyses which were independent of secular changes in the background population occurring over the same period.

### Infant Survival

Figure 2 shows Kaplan-Meier cumulative survival estimates with 95% confidence intervals for the first 365 days of life, stratified by generation. Figure 3 describes hazard ratios estimating differences in survival in the first 365 days of life by generation, and further stratified by whether the mother or father was the descendent of a longevity-enriched sibship. Figure 2 shows strong divergence of survival curves between G3 grandchildren and general population controls within the first 30 days of life, with parallel trends thereafter. However, strong separation was not observed between survival curves when comparing G4 great-grandchildren to their matched controls.

**Figure 2.**
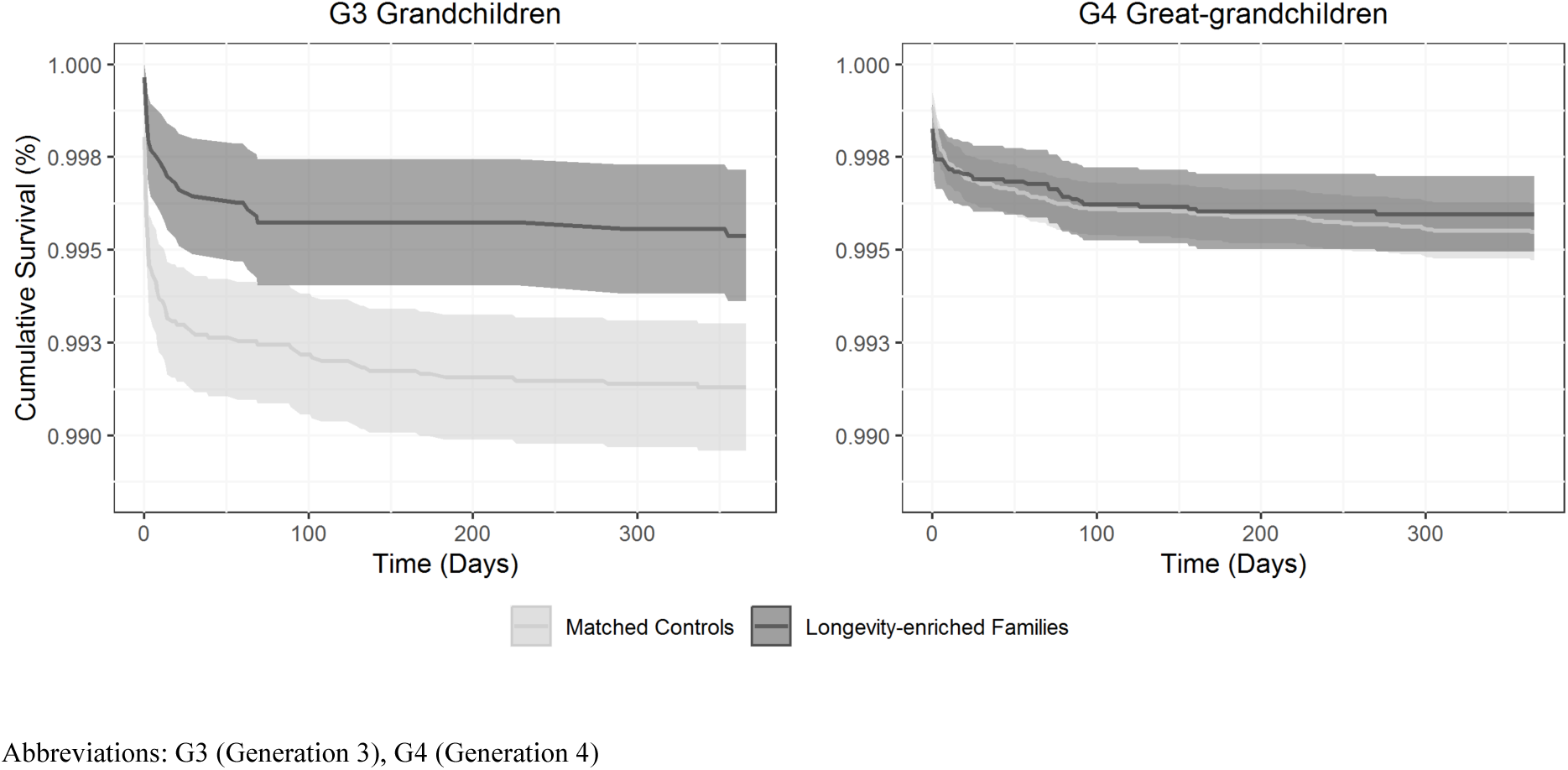
Kaplan-Meier cumulative survival estimates (95% CI) for G3 grandchildren and G4 great-grandchildren of longevity-enriched sibships and matched controls during the infant period

**Figure 3.**
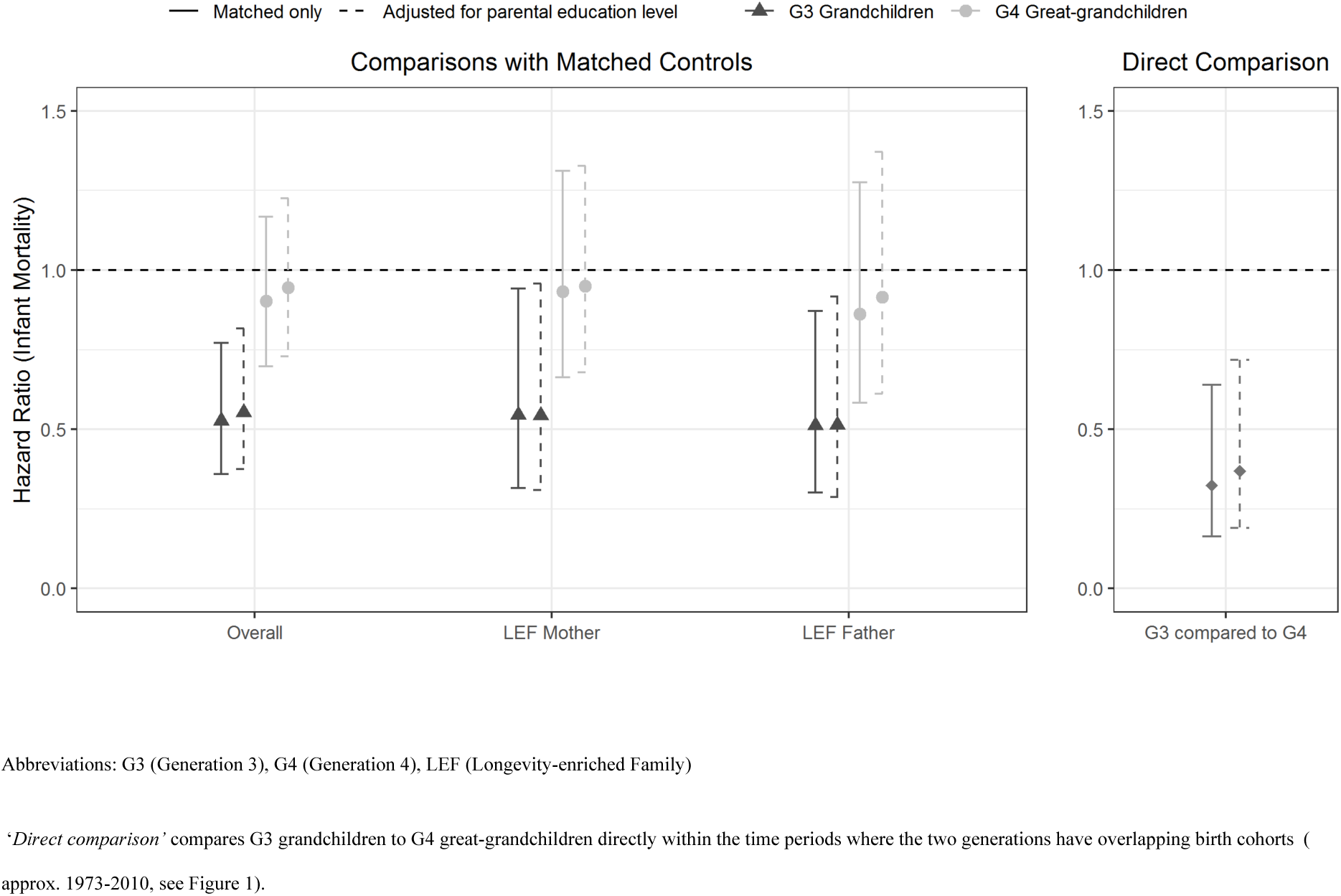
Cox regression analyses of infant survival comparing G3 grandchildren and G4 great-grandchildren of longevity-enriched sibships to matched controls

G3 grandchildren had approximately half the mortality of controls (HR = 0.53, 95% CI [0.36, 0.77]), and no evidence of differential effects dependent on LEF status of the mother or father (Interaction = 1.06, 95% CI [0.50, 2.28]). G4 great-grandchildren had a weaker association (HR = 0.90, 95% CI [0.70, 1.17], and similarly, no evidence of an interaction depending on maternal or paternal LEF status (HR = 1.08, 95% CI [0.64, 1.82]). All estimates were robust to further adjustment for highest attained parental level of education.

The Kaplan Meier survival curves in Figure 2 suggest that baseline survival was approximately equal in LEF grandchildren and great-grandchildren, possibly indicating that it was changes in the control group that was determining the dilution of the relative advantage in infant survival across generations. To test this hypothesis, we utilised the period of overlapping birth years between the two generations to perform an analysis directly comparing LEF grandchildren and great-grandchildren (see ‘Direct Comparison’ in Figure 3). Here, we observed a substantial reduction in mortality in G3 grandchildren compared to G4 great-grandchildren (HR = 0.32, 95% CI [0.16, 0.64]), independent of secular trends in the background population. This suggests there was a strong dilutionary effect in the infant survival advantage reflecting a loss of protective factors otherwise present in previous generations. Table S3.2 of the Supplementary Material describes a range of negative control sensitivity analyses that support this inference.

### Neonatal and Maternal Outcomes

Figure 4 displays odds ratios (ORs) from conditional logistic regression analyses of specific neonatal outcomes occurring in the first 28 days of life, or maternal outcomes throughout pregnancy, delivery and the puerperium. All ORs compared outcomes in live births of descendants of longevity-enriched sibships to those from the general population and were grouped by generation and adjustment for highest parental education level.

**Figure 4.**
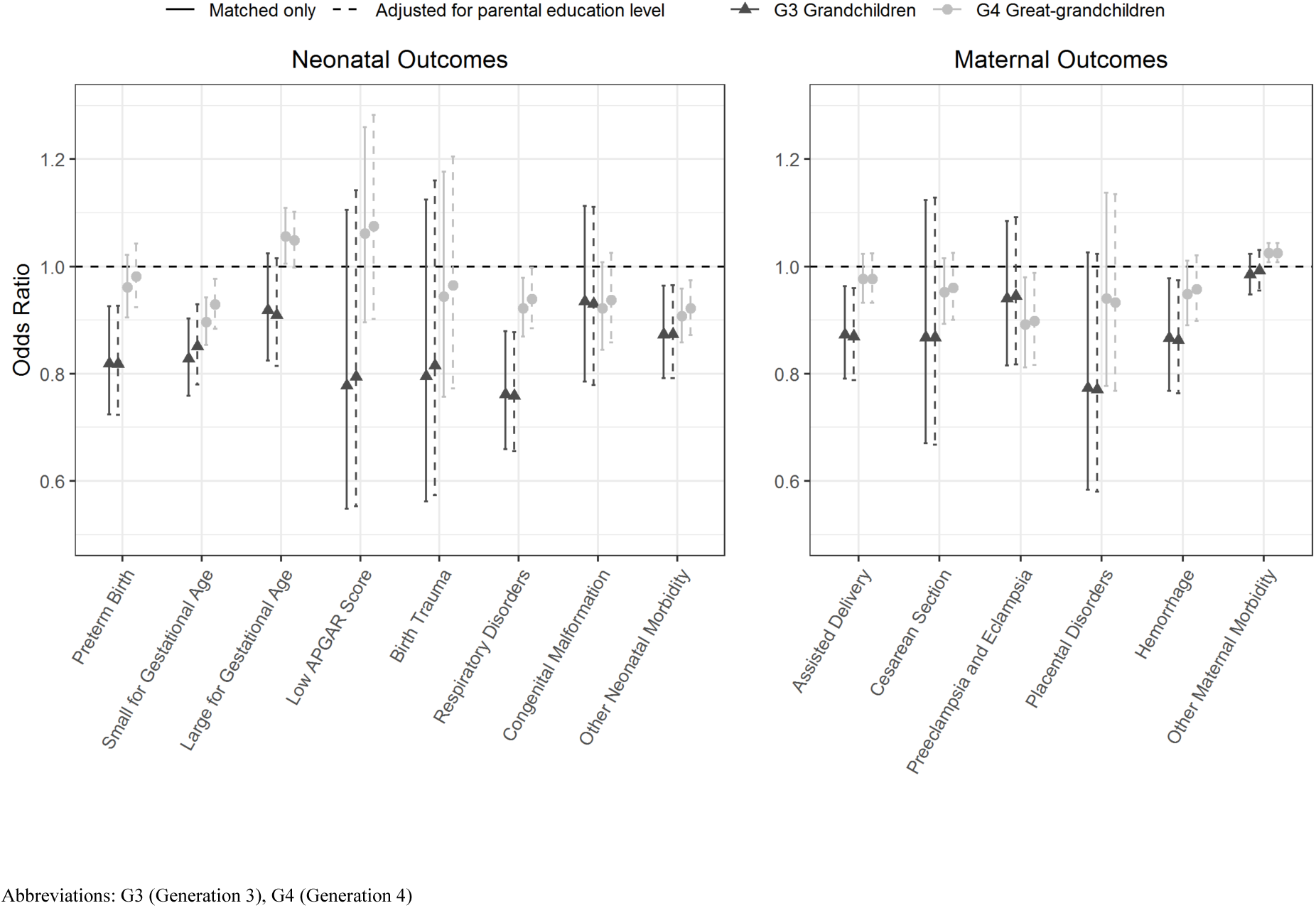
Conditional logistic regression analyses of neonatal and maternal outcomes in G3 grandchildren and G4 great-grandchildren of longevity-enriched sibships compared to matched controls

In G3 grandchildren, notable reductions were observed for preterm birth (OR = 0.82, 95% CI [0.72, 0.93]), small for gestational age (OR = 0.83, 95% CI [0.76, 0.90]), neonatal respiratory disorders (OR = 0.77, 95% CI [0.67, 0.88]), other neonatal morbidity (OR = 0.87, 95% CI [0.79, 0.96]), assisted delivery (OR = 0.87, 95% CI [0.79, 0.96]), and haemorrhage (OR = 0.87, 95% CI [0.77, 0.98]). However, all other associations were also negative in effect, implying a general trend towards reduced risk across a wide range of adverse birth outcomes. In G4 great-grandchildren, signals of risk reduction were weaker in magnitude and less consistent but were notable in the case of small for gestational age (OR = 0.90, 95% CI [0.85, 0.94]), neonatal respiratory disorders (OR = 0.92, 95% CI [0.87, 0.98]), and other neonatal morbidity (OR = 0.91, 95% CI [0.86, 0.96]). Minimal changes were observed after adjusting for parental education levels in both generations, suggesting a lack of confounding by parental socioeconomic status, as in the previous analyses of infant mortality.

After adjustment for multiple testing (n_outcomes_ = 14) and applying a 5% significance threshold, statistically significant differences were observed only in the following outcomes for G3 grandchildren (preterm birth, small for gestational age, and neonatal respiratory disorders) and G4 great-grandchildren (small for gestational age and other neonatal morbidity). Tables S2.3 and S2.4 of the Supplementary Material present estimates and adjusted p-values for all these analyses. Items S2.6-S2.9 of the Supplementary Material describe analyses of composite outcomes of neonatal and morbidity based on the individual measures included here.

### Maternal Smoking and Parental Education

Figure 5 describes the longitudinal completeness of data measuring maternal smoking behaviour in the Medical Birth Registry, and analyses comparing exposure to maternal smoking in mothers of LEF children compared to the general population. Data on maternal smoking was only available since 1991 but was also uniquely missing in 1997. The prevalence of maternal smoking was consistently lower in mothers of G3 grandchildren (OR = 0.80, 95% CI [0.69, 0.92]) and G4 great-grandchildren (OR = 0.75, 95% CI [0.71, 0.79]) of longevity-enriched sibships. These differences were attenuated after adjustment for parental education for both G3 grandchildren (OR = 0.83, 95% CI [0.72, 0.96]) and G4 great-grandchildren (OR = 0.84, 95% CI [0.80, 0.88]). Moreover, greater advantages in this behavioural trait were observed in mothers who were descendants of longevity-enriched sibships compared to mothers marrying into such families. Due to limited data in the births of third-generation grandchildren, we were not able to directly assess a mediating role of maternal smoking behaviour in their advantages in specific infant and neonatal outcomes.

**Figure 5.**
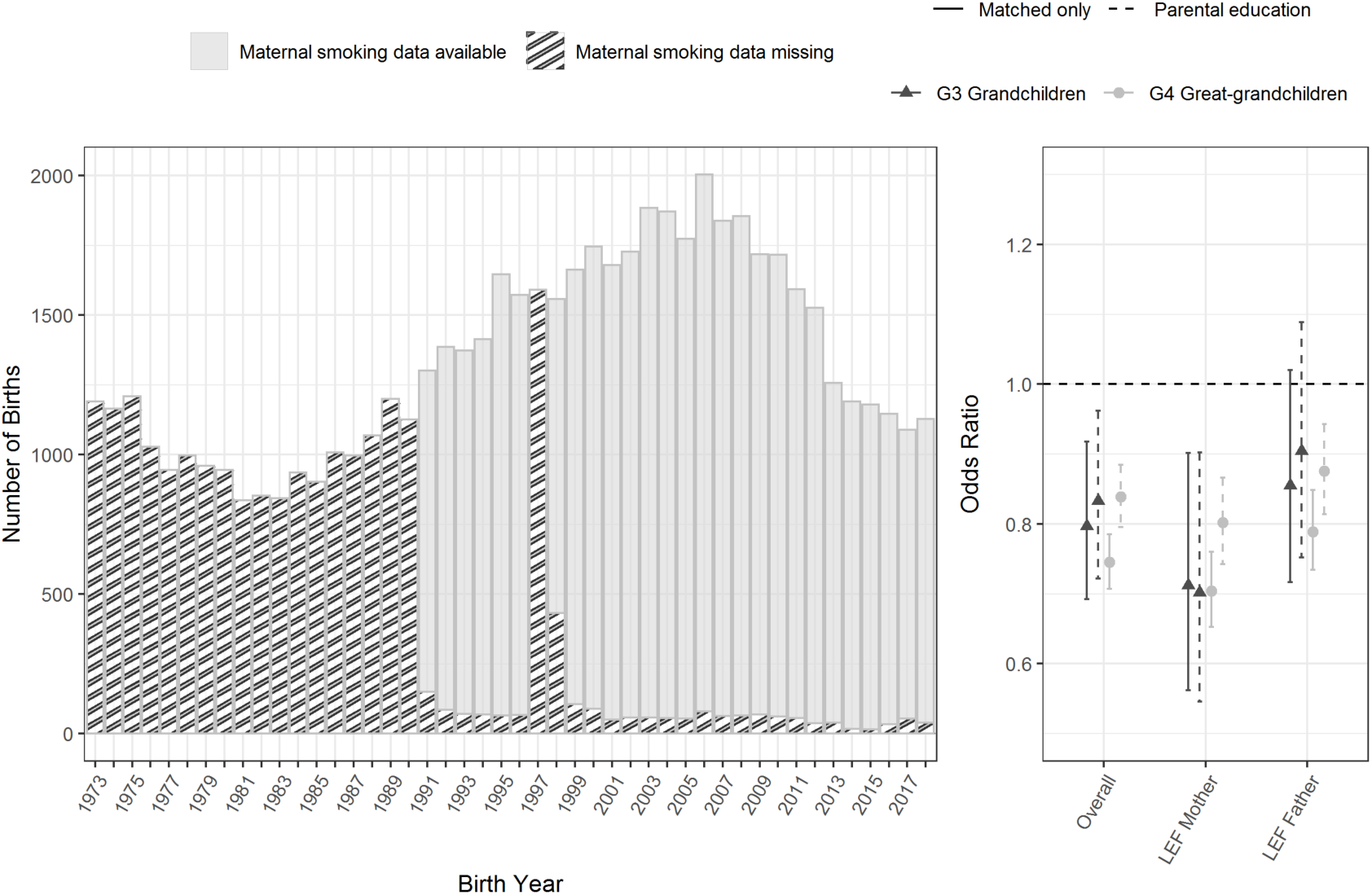
Completeness of maternal smoking data in the medical birth registry and conditional logistic regression analyses of exposure to maternal smoking in G3 grandchildren and G4 great-grandchildren of longevity-enriched sibships compared to matched controls Abbreviations: G3 (Generation 3), G4 (Generation 4), LEF (Longevity-enriched Family)

Figure 6 describes analyses of highest attained education levels of parents of G3 grandchildren and G4 great-grandchildren of longevity-enriched sibships compared to parents of matched controls. The left panel describes a multinomial logistic regression analysis of education categories compared to primary or lower secondary education. The right panel describes the overall estimates obtained from ordinal logistic regression models with a proportional odds assumption. Parents of both G3 grandchildren and G4 great-grandchildren were more likely to have higher levels of education across all categories compared to matched controls. Assuming proportional odds between the combinations of ordinal categories, parents of both G3 grandchildren (OR = 1.28, 95% CI [1.21, 1.36]) and G4 great-grandchildren (OR = 1.34, 95% CI [1.29, 1.39]) were more likely to be higher educated than parents of general population matched controls. When comparing both generations directly in their overlapping calendar periods, parents of G3 did not have different odds of having more education (OR = 0.95, 95% CI [0.86, 1.06]) than parents of G4, suggesting stability of this trait over successive generations. Tables S2.4 and S2.5 of the Supplementary Material contain all estimates from our analyses of exposure to maternal smoking and parental educational attainment.

**Figure 6.**
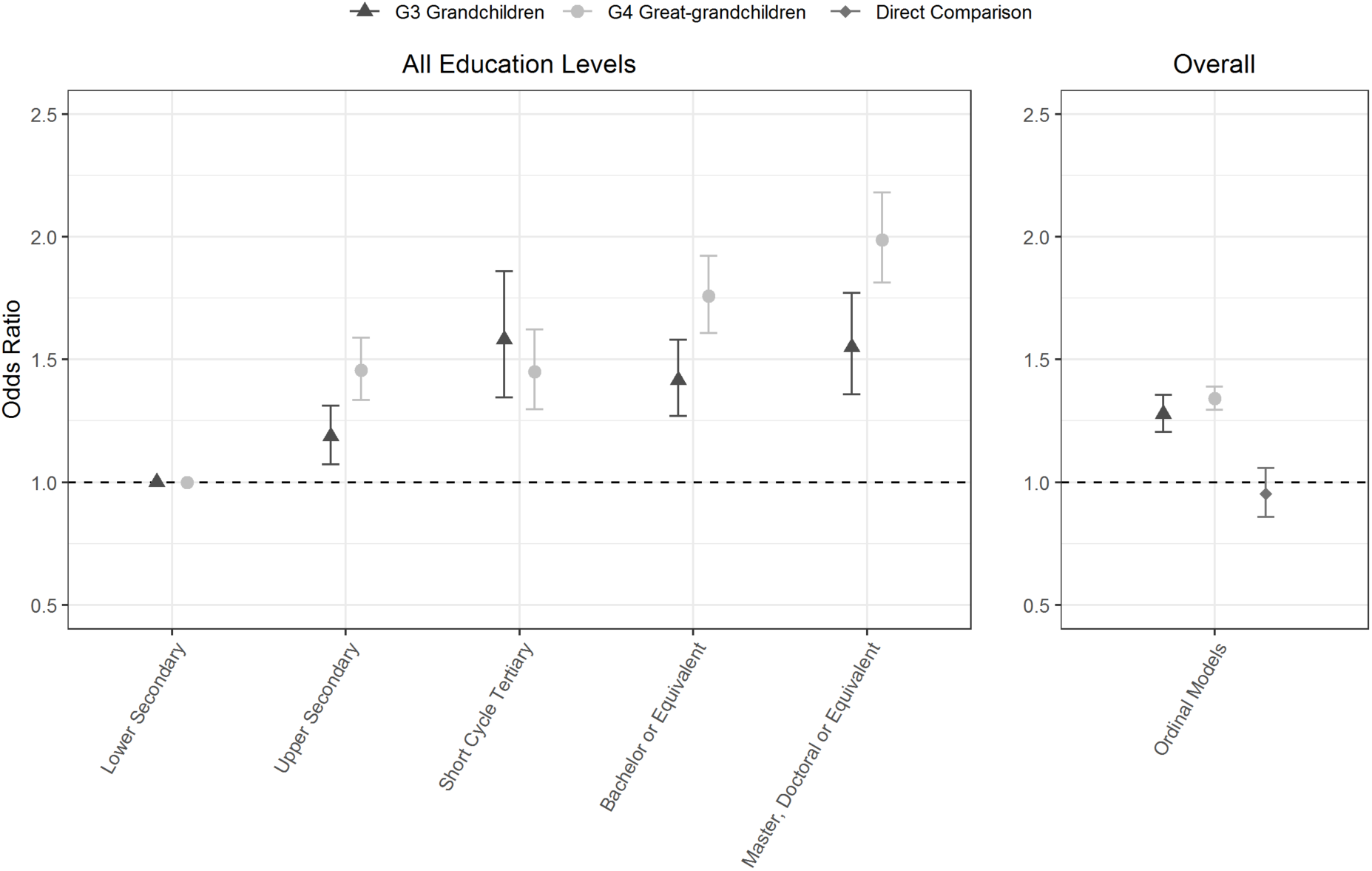
Multinomial and ordinal logistic regression analyses of highest attained education level in parents of G3 grandchildren and G4 great-grandchildren of longevity-enriched sibships compared to matched controls Abbreviations: G3 (Generation 3), G4 (Generation 4), LEF (Longevity-enriched Family) ‘Overall’ compares the odds of having any higher level of education in parents of longevity-enriched families to parents of matched controls, assuming proportional odds between the ordered categories. ‘*Direct comparison*’ in the ‘Overall’ figure compares G3 grandchildren to G4 great-grandchildren directly within the time periods where the two generations have overlapping birth cohorts (approx. 1973-2010, see Figure 1).

### Sensitivity Analyses

Our results were robust to further statistical adjustment for paternal country of birth and paternal age. Our findings on congenital anomalies were unchanged when considering longer diagnostic windows their detection. Analyses of continuous outcomes and more granular categorisations of key birth indicators (e.g., very/extremely small for gestational age) supported our primary study findings. Several outcomes with non-random missingness of data were detected and predisposed to a bias diluting the estimated protective effect against adverse outcomes in longevity-enriched families. For example, Apgar score measured after 5 minutes was disproportionately missing in those with an early neonatal death. Consistent findings were observed across analyses of composite neonatal and maternal morbidities, with various definitions including or excluding ‘other morbidity’ categories. Appending previously unmatched study participants to our data with weakly matched controls did not affect our main findings.

Lastly, negative control analyses of our ‘direct comparison’ methodology demonstrated that our results were not biased by secular improvements in neonatal outcomes and infant mortality when directly comparing between generations G3 and G4. This supported our interpretation that the exceptional infant phenotype is attenuated over successive generations in longevity-enriched families, and that this reflects a dilution of protective factors otherwise present in previous generations. Full reporting on our sensitivity analyses is available in sections S2 and S3 of the Supplementary Material.

## Discussion

Previous research has shown that the survival advantage in descendants of longevity-enriched sibships may manifest as early as the first year of life.^21^ However, little is known about what factors underlie this observation, other health manifestations beyond survival, or its transmission across generations. In this study we set out to explore the early life health trajectories of descendants of longevity-enriched sibships, utilising a multigenerational cohort and national birth registries in Denmark. We analysed patterns of infant survival and adverse birth outcomes in third-generation (G3) grandchildren and fourth-generation (G4) great-grandchildren descending from a proband generation of longevity-enriched sibships (G1). Compared to live births from the general population, G3 grandchildren demonstrated reductions in the risk of infant mortality and a range of adverse birth outcomes, independent of demographic and socioeconomic factors. However, these effects were strongly attenuated in G4 great-grandchildren with weaker and less consistent effects across all outcomes studied. The infant health advantage observed in our study manifested across a diverse range of outcomes, suggesting that survival differences were not driven by protection against a narrow range of conditions related to a particular aetiology.

Maternal and prenatal care in Denmark has changed considerably over the observation period included in our study.^32^ Such changes may have had differential effects depending on whether a child was born into a family with lower rather than exceptional health at baseline. Overlapping birth cohorts between successive generations in our study allowed us to directly assess the intergenerational transmission of the observed survival advantages in restricted samples, without reference to the general population. When comparing G3 grandchildren to G4 great-grandchildren within their overlapping birth cohorts (approx. birth years 1970 – 2010) we observed reduced mortality similar to the difference observed when comparing to contemporary matched controls in unrestricted samples. This methodology was also able to accurately capture expected patterns of survival when applied in a range of negative control comparisons involving G3 and G4 matched controls (Supplementary Material S3.1 and S3.2). For example, G4 great-grandchildren were not different in survival compared to G3 matched controls, nor were G3 controls different to G4 controls. The dilution of the exceptional phenotype observed in our study was thus independent of secular trends in the background population and reflected a partial loss of protective factors otherwise present in previous generations in our cohort of longevity-enriched families.

Maternal smoking is a well-established risk factor for adverse birth outcomes in offspring, as well as pathological sequalae in later life.^29^ In our cohorts, we observed a lower rate of smoking during pregnancy in mothers of both G3 grandchildren and G4 great-grandchildren. This finding supports previous research by our group implicating behavioural factors in the mechanisms underlying the familial aggregation of exceptional health and survival, including reduced risk of tobacco-related cancers in G2 offspring and G3 grandchildren.^20,21^ Interestingly, the smoking advantage was present to the same degree in both generations after adjustment for parental education levels, suggesting stability of this behavioural trait over successive generations. Furthermore, we observed a greater advantage in mothers who were a descendent of a longevity-enriched sibship, as opposed to mothers who married into such families. Both were superior to general population controls, suggesting a role for shared environmental effects, assortative mating, and or indirect genetic effects in the aggregation of positive behavioural traits in longevity-enriched families.^33,34^

Spouses marrying into our cohort of longevity-enriched families exhibit survival that is less exceptional than their partners, but greater than the general population.^35^ This relatively smaller advantage likely contributes to the dilution of exceptional health and survival across successive generations.^14,36,37^ Thus, we tested the relative importance of cumulative maternal versus paternal factors in the transmission of the exceptional infant phenotype by comparing children with mothers versus fathers descending from a longevity-enriched family.^38^ Remarkably similar patterns of infant survival were observed in both groups. This finding interpreted in isolation may suggest either a negligible effect of *parental* environment factors relative to heritable factors, or an approximately equal role of both paternal and maternal environmental factors. Due to mixed evidence across generations, we could not rule out modest differences in the importance of maternal and paternal environmental factors in transmission for other neonatal outcomes (see Supplementary Material S2.6 – S2.9).

Infant health outcomes have a proximal and limited range of determinants and depend less on complex interactions between factors later on in the life course, such as behaviour.^39^ These proximal factors include strictly biological genetic and epigenetic factors directly transmitted from parent to offspring, as well as environmentally mediated factors relating to foetal nutrition, maternal behaviour and health during pregnancy, and the early postnatal environment. In our cohort, we observed an infant health and survival advantage declining across successive generations with increasing distance from the longevity-enriched proband sibships. This pattern likely reflected a partial loss of protective factors otherwise present in previous generations. We also observed remarkably consistent advantages in behavioural and socioeconomic indicators in the parents of both G3 grandchildren and G4 great-grandchildren. This suggests that maternal smoking and education level, as well as other factors proxied by their measurement, were not primary drivers of the exceptional infant phenotype observed in G3 grandchildren.

Lastly, we observed strong similarity in maternal and paternal lines of transmission, despite the known vulnerability of the foetus to maternal nutrition and physiology.^38^ This is likely best interpreted as minimising the role of environmentally mediated factors in the mechanisms underlying the exceptional infant phenotype in this study.

These patterns in our view strongly implicate heritable genetic and/or epigenetic factors in the transmission of the exceptional infant phenotype in our cohort of longevity-enriched families. This discussion pertains to the developmental mechanisms driving the infant health advantage specifically, and do not preclude an increasingly important role for behaviourally mediated effects manifesting directly in childhood and beyond, or indirectly through the familial environment. Indeed, research by our group has identified patterns of disease risk and family stability suggesting behaviour plays a key role in the aggregation of exceptional health and survival in longevity- enriched families.^20,21^ Although we were unable to observe G2 offspring and G1 sibships in their developmental periods, we have previously shown a similar dilution of adult mortality across generations G2 and G3.^21^ It is likely that part of this dilution may also be due to a partial loss of heritable genetic and or epigenetic protective factors. However, the relationship between these complex factors and how they interact throughout the life course in the familial aggregation of longevity is currently unknown. An important avenue for future research will be to assess the health and survival trajectories of G4 great-grandchildren and G3 grandchildren throughout mid- and late-life respectively.

The idea that environmental conditions in early life can directly influence health and functioning later in life has been subject to investigation and embraced by a range of scientific disciplines. Studies of prospective cohorts, experimental manipulation in model organisms, and quasi-random environmental shocks in early life (e.g. famine, business cycles, and natural disasters) have coalesced to form the framework for the Developmental Origins of Health and Disease (DOHaD) hypothesis.^40^ Maladaptive responses to developmental cues, generally meant to preserve genotypic variation in the face of transitory environmental changes, are likely central to this phenomenon.^41^ Developmental effects have been implicated in a range of adult diseases including cardiovascular and metabolic disease, cancer, and neuropsychiatric conditions.^42^ Epigenetic mechanisms are a suspected to play an important role in observations of multigenerational and transgenerational effects arising from developmental exposures in particular.^43^

Results from the present study and previous research by our group point to a highly favourable profile of parental risk factors which are commonly implicated in developmental mechanisms of intergenerational disease transmission.^21^ For example, G2 offspring and G3 grandchildren in our cohort exhibit lowered risk of disease incidence and mortality due to mental and behavioural disorders, cardiovascular and endocrine diseases, and infections during adulthood.^21^ Social stress, substance abuse, infections, and cardiometabolic disease have all been established as probable causative factors within the DOHaD framework.^44–47^ Interestingly, measurable changes in common birth outcomes are not necessary for long-term physiological changes in response to an adverse prenatal and early postnatal conditions.^48^ From these perspectives, our findings suggest that the familial aggregation of exceptional longevity may have developmental origins as early as the perinatal period.

To our knowledge, this is the first study to comprehensively assess early life health trajectories associated with familial aggregation of longevity.^49,50^ Such efforts have previously not been feasible due to a lack of prospective cohorts or clinical birth registries with sufficient history of follow-up.^15^ In our view, the patterns of transmission between generations G3 and G4 observed in our study predict certain trends in early life health trajectories of previous generations, which we were unable to directly observe. Specifically, it is likely that the phenotype observed in G3 grandchildren in our cohort may represent a conservative portrayal of the G2 offspring and G1 proband generations in their unobserved perinatal and infant periods. In this way, extrapolations of our findings could aid in developing our scientific knowledge of early life health trajectories more directly associated with phenotypic longevity. However, it remains unknown to what extent such exceptional infant phenotypes predict familial aggregation of longevity within a single generation, and this would be an important avenue for future research, once methodologically feasible to do so.

This study had several important strengths and limitations. Due to the nationwide scope our study and the population-based ascertainment of our cohort, there was minimal attrition in the follow-up of descendants of the proband longevity-enriched sibships. Our use of national birth and patient registries permitted analyses of clinically measured outcomes over successive generations and facilitated comparisons with the general Danish population. However, data on maternal smoking during pregnancy was missing for a large portion of the G3 generation. Thus, we could not perform reliable analyses assessing its mediating role across a variety of statistically important differences. Smoking data for both generations was only available with binary classification, and this may have missed differences in more granular measures, including trajectories of smoking cessation across the trimesters of pregnancy. Lastly, our inferences based on patterns of maternal smoking and behavioural indicators over successive generations may be subject to residual confounding. However, given the stability of these very broad proxies, it is unlikely that more granular measurements of behaviour and socioeconomic status could explain the dilution of the exceptional infant phenotype over generations.

## Conclusion

Descendants of longevity-enriched sibships exhibit a broad health advantage manifesting as early the perinatal period. The infant survival advantage in these families was driven by protection against a diverse range of adverse birth outcomes, and independent of socioeconomic and behavioural factors, as well as maternal or paternal lines of transmission. Health and survival effects were strongly diluted over successive generations, despite persistent behavioural and socioeconomic advantages. Our findings suggest that exceptional health and survival may have early developmental components and implicate heritable genetic and or epigenetic factors in their transmission.

## Data Availability

Data for this research was obtained on a per-project basis in liaison with a government agency in Denmark (Statistics Denmark) and there are strict restrictions on its use and sharing. The data cannot be deposited in a public database and exports of summary data is only allowed as material for direct use in a scientific publication.

## Abbreviations

HR: hazard ratio
OR: odds ratio
LEF: longevity-enriched family
LLFS: Long Life Family Study
G3: Generation 3
G4: Generation 4

## Contributions

KC, MTK, and PLS conceived the study idea. MTK, KC, MFF, MW and MP designed the study. MTK and DAP obtained and pre-processed the study data. MTK performed the data analysis and takes full responsibility for the integrity of the results. MTK and KC wrote the initial manuscript. All authors worked on subsequent iterations of the manuscript and contributed intellectual content. All authors approved the final manuscript.

## Ethical Approval

The study has been ethically approved by The Regional Scientific Ethical Committees for Southern Denmark (S-VF-20030227), The Danish Data Protection Agency (J.nr. 2008-41-1753), and University of Southern Denmark, Research & Innovation Organisation (J.nr. 10.635).

## Transparency

The manuscript’s guarantor (MTK) affirms that this manuscript is an honest, accurate and transparent account of the study being reported; that no important aspects of the study have been omitted; and that any discrepancies from the study as originally planned have been explained.

