## Supplementary Material for "Developmental origins of exceptional health and survival: A four-generation family cohort study"

### Supplementary Appendix for ‘Developmental origins of exceptional health and survival: a four-generation family cohort study’

Matthew Thomas Keys[1]*; Dorthe Almind Pedersen[1]; Pernille Stammen Larsen[1]; Alexander Kulminski[2]; Mary F. Feitosa[3]; Mary Wojczynski[3]; Michael Province[3]; Kaare Christensen[1,4]

[1]Department of Epidemiology, Biostatistics, and Biodemography, University of Southern Denmark, Odense, Denmark

[2]Social Science Research Institute, Duke University, Durham, North Carolina, USA

[3]Department of Genetics, Washington University School of Medicine, St Louis, Missouri, USA

[4]Danish Ageing Research Centre, University of Southern Denmark, Odense, Denmark

### Contents

1. Methods and Materials
   - ***Figure S1.1.*** Structure of Danish Longevity-Enriched Families (LEFs)
   - ***Table S1.2.*** ICD-8 and ICD-10 codes used to define maternal and neonatal outcomes

2. Additional results from main analyses
   - ***Figure S2.1.*** Derivation of study population from source population of G3 grandchildren and G4 great-grandchildren (corresponding to **Table 1** in article)
   - ***Table S2.2.*** Regression results for analyses of mortality in G3 grandchildren and G4 great-grandchildren (corresponding to **Figure 3** in article)
   - ***Table S2.3.*** Regression results for analyses of neonatal morbidities in G3 grandchildren and G4 great-grandchildren, with adjusted p-values (corresponding to **Figure 4** in article)
   - ***Table S2.4.*** Regression results for analyses of maternal morbidities in mothers of G3 grandchildren and G4 great-grandchildren, with adjusted p-values (corresponding to **Figure 4** in article)
   - ***Table S2.4.*** Regression results for analyses of exposure to maternal smoking in G3 grandchildren and G4 great-grandchildren (corresponding to **Figure 5** in article)
   - ***Table S2.5.*** Regression results for analyses of highest attained education level in parents of G3 grandchildren and G4 great-grandchildren (corresponding to **Figure 6** in article)
   - ***Figure S2.6.*** Regression results for analyses of composite neonatal and maternal morbidity in G3 grandchildren and G4 great-grandchildren and their parents (definition 1)
   - ***Figure S2.7.*** Regression results for analyses of composite neonatal and maternal morbidity in G3 grandchildren and G4 great-grandchildren and their parents (definition 2)
   - ***Table S2.8.*** Regression results for analyses of composite neonatal and maternal morbidity in G3 grandchildren and G4 great-grandchildren and their parents (definition 1)
   - ***Table S2.9.*** Regression results for analyses of composite neonatal and maternal morbidity in G3 grandchildren and G4 great-grandchildren and their parents (definition 2)

3. Sensitivity and Robustness Analyses
   - ***Figure S3.1.*** Regression results for negative control analyses using overlapping generations and matched controls
   - ***Table S3.2.*** Regression results for negative control analyses using overlapping generations and matched controls
   - ***Table S3*.3.** Regression results for analyses with varying windows of opportunity for the diagnosis of congenital malformations
   - ***Table S3.4.*** Regression results for analyses of mortality when adjusting for paternal immigration status and age at birth
   - ***Table S3.5.*** Regression results for analyses assessing other outcomes using weight at birth

- ***Table S3.6.*** Regression results for analyses of key neonatal indicators as continuous rather than dichotomized outcomes
- ***Table S3.7.*** Regression analyses for analyses of mortality including weakly matched sets who were previously unable to be matched according to the full matching criteria
- ***Table S3.8.*** Missingness of data in key outcomes derived from the Medical Birth Registry stratified by levels of censoring variables
- ***Table S3.7.*** Missingness of data in key outcomes derived from the Medical Birth Registry stratified by levels of censoring variables

#### 1. Methods and Materials

***Figure S1.1.*** Structure of Danish Longevity-Enriched Families (LEFs)

Please see the supplementary appendix for Aging Cell 'Mechanisms underlying the familial aggregation of exceptional health and survival: A three-generation cohort study' for further details on the construction of our cohort of longevity-enriched families.

Paper:
https://onlinelibrary.wiley.com/doi/full/10.1111/acel.13228

Supplementary material:
https://onlinelibrary.wiley.com/action/downloadSupplement?doi=10.1111%2Facel.13228&file=acel13228-sup-0001-AppendixS1.docx

***Table S1.2.*** ICD-8 and ICD-10 codes used to define maternal and neonatal outcomes

| Type | Outcome | ICD Level | ICD CODES | Notes |
|---|---|---|---|---|
| | Preterm Birth | NA | NA | Calculated from gestational age variable |
| | Small for gestational age | NA | NA | Calculated from gestational age and birth weight variables |
| | Large for gestational age | NA | NA | Calculated from gestational age and birth weight variables |
| | Low APGAR Score | NA | NA | Calculated from gestational age and birth weight variables |
| | Birth Trauma | ICD-8 | 764-768, 772 | All codes inclusive |
| | | ICD-10 | P10-P15 | All codes inclusive |
| **Neonatal** | Neonatal respiratory disorders | ICD-8 | 763-768*, 776 | *With mention of hypoxia |
| | | ICD-10 | P20-P28 | All codes inclusive |
| | Congenital malformation | ICD-8 | 740 - 759 | All codes inclusive |
| | | ICD-10 | Q00 - Q99 | All codes inclusive |
| | Other neonatal morbidity | ICD-8 | 760-779**☆ | *All adverse neonatal codes not used elsewhere, ☆with exceptions |
| | | ICD-10 | P00-P96**☆ | *All adverse neonatal codes not used elsewhere, ☆with exceptions |

| | | | | |
|---|---|---|---|---|
| | Assisted delivery | ICD-8 | 650-662* | *With mention of forceps, vacuum extractor or other instrumental methods |
| | | ICD-10 | O81, O83, O84* | *With mention of forceps or vacuum extractor |
| | Cesarean section | ICD-8 | 650-662* | *With mention of cesarean section |
| | | ICD-10 | O82, O84* | *With mention of cesarean section |
| **Maternal** | Preeclampsia and eclampsia | ICD-8 | 637 | All codes inclusive |
| | | ICD-10 | O11, O14, O15 | All codes inclusive |
| | Placental disorders | ICD-8 | 632*, 634*, 651 | *With mention of placenta☆ |
| | | ICD-10 | O43-O45 | All codes inclusive☆ |
| | Hemorrhage | ICD-8 | 632, 651, 653 | All codes inclusive, excluding postpartum hemorrhage☆ |
| | | ICD-10 | O20, O44*, O45*, O46, O67 | *With mention of hemorrhage, excluding postpartum hemorrhage☆ |
| | Other maternal morbidity | ICD-8 | 630-678**☆ | **All adverse maternal codes not used elsewhere, ☆with exceptions |
| | | ICD-10 | O00-O99**☆ | **All adverse maternal codes not used elsewhere, ☆with exceptions |

☆Please see written exceptions on next page.

☆**Exceptions and other explanations**

See below for which codes were excluded from our measures of 'other neonatal morbidity' and 'other maternal morbidity'. *Explanations* follow in italics.

Exclusion from **other neonatal morbidity**:

- ICD-8:
    - Foetal death of unknown cause (779) – *defined elsewhere*
    - Termination of pregnancy (773) – *not relevant to live births*
- ICD-10:
    - Disorders related to length of gestation and foetal growth (P05-P08) – *defined elsewhere*
    - Foetal death of unknown cause (P95) – *defined elsewhere*

Exclusion from hemorrhage:

- ICD-8
    - Delivery complicated by retained placenta (652) – *not directly relevant to neonatal health, included in 'other maternal morbidity'*
    - Delivery complicated by other postpartum hemorrhage (653) – *not directly relevant to neonatal health, included in 'other maternal morbidity'*
- ICD-10
    - Foetal death of unknown cause (O72) – *defined elsewhere*

Exclusion from placental disorders:

- ICD-8
    - Retained placenta (xxx) – *not directly relevant to neonatal health, included in 'other maternal morbidity'*
- ICD-10
    - Retained placenta (xxx) – *not directly relevant to neonatal health, included in 'other maternal morbidity'*

Exclusion from **other maternal morbidity**:

- ICD-8:
    - Ectopic pregnancy (631) – *not relevant to live births*
    - Abortion (640-645) – *not relevant to live births*
    - Delivery without mention of complication (6500, 6501, 6509) – *not adverse*
    - Anaesthetic death in uncomplicated delivery (662) – *maternal mortality not assessed*
    - Maternal care (Y60-Y69) – *not adverse*
    - Healthy live-born infants (Y80-Y89) – *not adverse*
- ICD-10:
    - Pregnancy with abortive outcome (O00-O08) – *not relevant to live births*

- Maternal care or care for suspected abnormality (O28, O30, O32-36)) – *not adverse*
- Multiple gestation (O30) – *not adverse*
- False labour (O47) – *not adverse*
- Preterm and long labour and delivery (O60, O48, O63) – *defined neonatally*
- Spontaneous Delivery (O80, O840) – *not adverse*
- Obstetric death (O95-O97) – *maternal mortality not assessed*

Important considerations:

1. Several disease groups contain codes used in other disease groups. This is because of the hierarchical structure of some of the codes in either in ICD-8 or ICD-10. For example, ICD-8 code 651 describes 'Delivery complication by placenta previa or antepartum haemorrhage'. But its subcodes (e.g. 651.0 – 651.8 refer to types of assisted delivery or caesarean section). In this case, 650 codes could appear simultaneously for groups for maternal haemorrhage, assisted delivery and caesarean section. There are several more examples like this.
2. ICD-9 was not employed in Denmark, and so we only use ICD-8 (1973 – 1993) and ICD-10 (1994 - 2018) codes in this study.

#### 2. Additional results from main analyses

***Figure S2.1.*** Derivation of study population from source population of G3 grandchildren and G4 great-grandchildren (corresponding to **Table 1** in article)

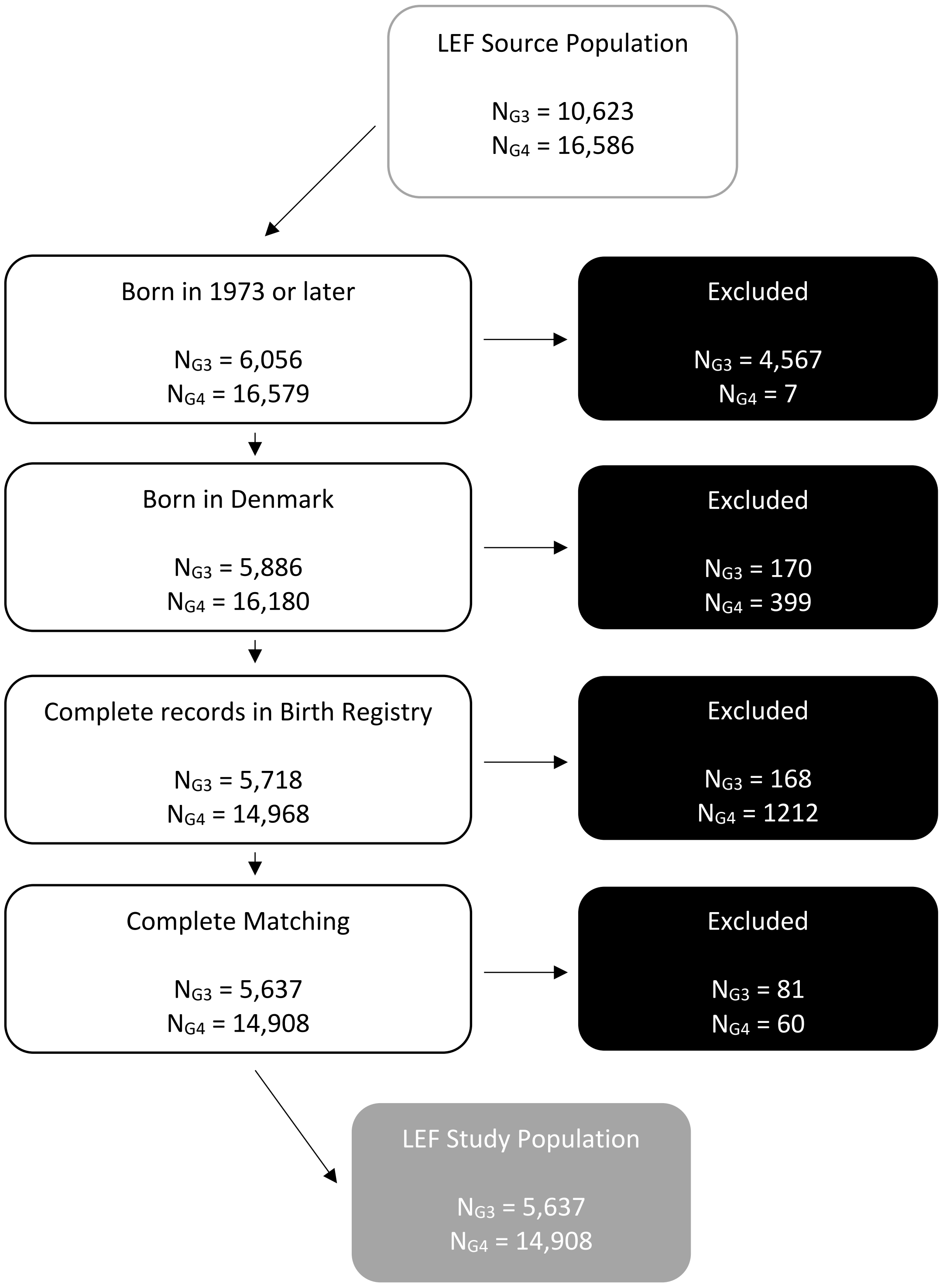

***Table S2.2.*** Regression results for analyses of mortality in G3 grandchildren and G4 great-grandchildren (corresponding to **Figure 3** in article)

| Generation | Model | Adjustment | HR | 95% CI | p |
|---|---|---|---|---|---|
| Grandchildren | LEF vs Matched Controls | Matched only | 0.53 | [0.36, 0.77] | 0.001 |
| | | Parental education | 0.55 | [0.37, 0.82] | 0.003 |
| | LEF Mother Subset | Matched only | 0.55 | [0.32, 0.94] | 0.030 |
| | | Parental education | 0.54 | [0.31, 0.96] | 0.035 |
| | LEF Father Subset | Matched only | 0.51 | [0.29, 0.92] | 0.014 |
| | | Parental education | 0.51 | [0.29, 0.92] | 0.024 |
| | Interaction | Matched only | 1.06 | [0.50, 2.28] | 0.872 |
| | | Parental education | 1.11 | [0.51, 2.44] | 0.793 |
| Great-grandchildren | LEF vs Matched Controls | Matched only | 0.90 | [0.70, 1.17] | 0.433 |
| | | Parental education | 0.95 | [0.73, 1.23] | 0.672 |
| | LEF Mother Subset | Matched only | 0.93 | [0.66, 1.31] | 0.691 |
| | | Parental education | 0.95 | [0.68, 1.33] | 0.764 |
| | LEF Father Subset | Matched only | 0.86 | [0.58, 1.28] | 0.458 |
| | | Parental education | 0.92 | [0.61, 1.37] | 0.670 |
| | Interaction | Matched only | 1.08 | [0.64, 1.82] | 0.764 |
| | | Parental education | 1.02 | [0.60, 1.73] | 0.952 |
| Direct Comparison | Grandchildren vs great-grandchildren | Matched only | 0.32 | [0.16, 0.64] | 0.001 |
| | | Parental education | 0.37 | [0.19, 0.72] | 0.003 |

***Table S2.3.*** Regression results for analyses of neonatal morbidities in G3 grandchildren and G4 great-grandchildren, with adjusted p-values (corresponding to **Figure 4** in article)

| Generation | Neonatal Outcome | Adjustment | OR | 95% CI | p | $p_{adjusted}$ |
|---|---|---|---|---|---|---|
| G3 | Preterm Birth | Matched only | 0.82 | [0.72, 0.93] | 0.001 | 0.02 |
| | | Parental education | 0.82 | [0.72, 0.93] | 0.002 | 0.02 |
| | Small for Gestational Age | Matched only | 0.83 | [0.76, 0.9] | < 0.001 | < 0.001 |
| | | Parental education | 0.85 | [0.78, 0.93] | < 0.001 | 0.004 |
| | Large for Gestational Age | Matched only | 0.92 | [0.82, 1.02] | 0.13 | 0.45 |
| | | Parental education | 0.91 | [0.81, 1.02] | 0.09 | 0.53 |
| | Low APGAR Score | Matched only | 0.78 | [0.55, 1.11] | 0.16 | 0.45 |
| | | Parental education | 0.79 | [0.55, 1.14] | 0.21 | 0.64 |
| | Birth Trauma | Matched only | 0.79 | [0.56, 1.12] | 0.19 | 0.45 |
| | | Parental education | 0.82 | [0.57, 1.16] | 0.26 | 0.66 |
| | Respiratory Disorders | Matched only | 0.76 | [0.66, 0.88] | < 0.001 | 0.003 |
| | | Parental education | 0.76 | [0.66, 0.88] | < 0.001 | 0.003 |
| | Congenital Malformation | Matched only | 0.93 | [0.79, 1.11] | 0.45 | 0.45 |
| | | Parental education | 0.93 | [0.78, 1.11] | 0.42 | 0.68 |
| | Other Neonatal Morbidity | Matched only | 0.87 | [0.79, 0.96] | 0.007 | 0.07 |
| | | Parental education | 0.87 | [0.79, 0.97] | 0.008 | 0.08 |
| G4 | Preterm Birth | Matched only | 0.96 | [0.91, 1.02] | 0.2 | 0.61 |
| | | Parental education | 0.98 | [0.92, 1.04] | 0.55 | 0.75 |
| | Small for Gestational Age | Matched only | 0.90 | [0.85, 0.94] | < 0.001 | < 0.001 |
| | | Parental education | 0.93 | [0.88, 0.98] | 0.004 | 0.05 |
| | Large for Gestational Age | Matched only | 1.06 | [1, 1.11] | 0.03 | 0.29 |
| | | Parental education | 1.05 | [1, 1.1] | 0.06 | 0.51 |
| | Low APGAR Score | Matched only | 1.06 | [0.9, 1.26] | 0.49 | 0.61 |
| | | Parental education | 1.08 | [0.9, 1.28] | 0.42 | 0.75 |
| | Birth Trauma | Matched only | 0.94 | [0.76, 1.18] | 0.61 | 0.61 |
| | | Parental education | 0.96 | [0.77, 1.21] | 0.75 | 0.75 |
| | Respiratory Disorders | Matched only | 0.92 | [0.87, 0.98] | 0.008 | 0.09 |
| | | Parental education | 0.94 | [0.88, 1] | 0.04 | 0.37 |
| | Congenital Malformation | Matched only | 0.92 | [0.84, 1.01] | 0.07 | 0.47 |
| | | Parental education | 0.94 | [0.86, 1.02] | 0.16 | 0.68 |
| | Other Neonatal Morbidity | Matched only | 0.91 | [0.86, 0.96] | < 0.001 | 0.007 |
| | | Parental education | 0.92 | [0.87, 0.97] | 0.004 | 0.05 |

***Table S2.4.*** Regression results for analyses of maternal morbidities in mothers of G3 grandchildren and G4 great-grandchildren, with adjusted p-values (corresponding to **Figure 4** in article)

| Generation | Maternal Outcome | Adjustment | OR | 95% CI | p | $p_{adjusted}$ |
|---|---|---|---|---|---|---|
| G3 | Assisted Delivery | Matched only | 0.87 | [0.79, 0.96] | 0.007 | 0.07 |
| | | Parental education | 0.87 | [0.79, 0.96] | 0.005 | 0.05 |
| | Cesarean Section | Matched only | 0.87 | [0.67, 1.12] | 0.28 | 0.45 |
| | | Parental education | 0.87 | [0.67, 1.13] | 0.29 | 0.66 |
| | Preeclampsia and Eclampsia | Matched only | 0.94 | [0.81, 1.08] | 0.4 | 0.45 |
| | | Parental education | 0.94 | [0.82, 1.09] | 0.44 | 0.68 |
| | Placental Disorders | Matched only | 0.77 | [0.58, 1.03] | 0.07 | 0.45 |
| | | Parental education | 0.77 | [0.58, 1.02] | 0.07 | 0.5 |
| | Hemorrhage | Matched only | 0.87 | [0.77, 0.98] | 0.02 | 0.18 |
| | | Parental education | 0.86 | [0.76, 0.97] | 0.02 | 0.15 |
| | Other Maternal Morbidity | Matched only | 0.98 | [0.95, 1.02] | 0.43 | 0.45 |
| | | Parental education | 0.99 | [0.95, 1.03] | 0.68 | 0.68 |
| G4 | Assisted Delivery | Matched only | 0.98 | [0.93, 1.02] | 0.33 | 0.61 |
| | | Parental education | 0.98 | [0.93, 1.02] | 0.33 | 0.75 |
| | Cesarean Section | Matched only | 0.95 | [0.89, 1.02] | 0.13 | 0.61 |
| | | Parental education | 0.96 | [0.9, 1.03] | 0.22 | 0.73 |
| | Preeclampsia and Eclampsia | Matched only | 0.89 | [0.81, 0.98] | 0.02 | 0.16 |
| | | Parental education | 0.90 | [0.82, 0.99] | 0.03 | 0.26 |
| | Placental Disorders | Matched only | 0.94 | [0.78, 1.14] | 0.53 | 0.61 |
| | | Parental education | 0.93 | [0.77, 1.13] | 0.49 | 0.75 |
| | Hemorrhage | Matched only | 0.95 | [0.89, 1.01] | 0.1 | 0.6 |
| | | Parental education | 0.96 | [0.9, 1.02] | 0.18 | 0.73 |
| | Other Maternal Morbidity | Matched only | 1.03 | [1.01, 1.04] | 0.004 | 0.05 |
| | | Parental education | 1.03 | [1.01, 1.04] | 0.004 | 0.05 |

***Table S2.4.*** Regression results for analyses of exposure to maternal smoking in G3 grandchildren and G4 great-grandchildren (corresponding to **Figure 5** in article)

| Generation | Outcome | Adjustment | Odds Ratio | 95% CI | p |
|---|---|---|---|---|---|
| G3 | Overall | Matched only | 0.80 | [0.69, 0.92] | 0.002 |
| | | Parental education | 0.83 | [0.72, 0.96] | 0.01 |
| | LEF Mother | Matched only | 0.71 | [0.56, 0.9] | 0.005 |
| | | Parental education | 0.70 | [0.55, 0.9] | 0.006 |
| | LEF Father | Matched only | 0.85 | [0.72, 1.02] | 0.08 |
| | | Parental education | 0.90 | [0.75, 1.09] | 0.29 |
| G4 | Overall | Matched only | 0.75 | [0.71, 0.79] | < 0.001 |
| | | Parental education | 0.84 | [0.8, 0.88] | < 0.001 |
| | LEF Mother | Matched only | 0.70 | [0.65, 0.76] | < 0.001 |
| | | Parental education | 0.80 | [0.74, 0.87] | < 0.001 |
| | LEF Father | Matched only | 0.79 | [0.73, 0.85] | < 0.001 |
| | | Parental education | 0.88 | [0.81, 0.94] | < 0.001 |

***Table S2.5.*** Regression results for analyses of highest attained education level in parents of G3 grandchildren and G4 great-grandchildren (corresponding to **Figure 6** in article)

| Regression Model | Generation | Education Level | Odds Ratio | CI |
|---|---|---|---|---|
| Multinomial | G3 | Lower Secondary | 1.00 | - |
| | | Upper Secondary | 1.19 | [1.07, 1.31] |
| | | Short Cycle Tertiary | 1.58 | [1.35, 1.86] |
| | | Bachelor or Equivalent | 1.42 | [1.27, 1.58] |
| | | Master, Doctoral or Equivalent | 1.55 | [1.36, 1.77] |
| | G4 | Lower Secondary | 1.00 | - |
| | | Upper Secondary | 1.46 | [1.33, 1.59] |
| | | Short Cycle Tertiary | 1.45 | [1.3, 1.62] |
| | | Bachelor or Equivalent | 1.76 | [1.61, 1.92] |
| | | Master, Doctoral or Equivalent | 1.99 | [1.81, 2.18] |
| Ordinal | G3 | Odds of a higher education level | 1.28 | [1.21, 1.36] |
| | G4 | | 1.34 | [1.29, 1.39] |
| | G3 vs G4 | | 0.95 | [0.86, 1.06] |

Items S2.6-S2.9 assess differences in overall neonatal and maternal morbidities. We employed two definitions for composite measures of neonatal and maternal morbidity based on the individual measures of morbidity included in our study (**Figure 4** of the article). What contrasts these two sets of definitions is the exclusion or inclusion of the 'other morbidity' categories. This was due to the difficulty of interpreting the 'other' category as they included all adverse ICD codes not included in the other categories, and capture highly diverse clinical phenomena in terms of both aetiology and severity.

**Neonatal morbidity definition 1:** Any occurrence of preterm birth, small for gestational age, large for gestational age, low APGAR score, birth trauma, respiratory disorders, or congenital malformation. *Excluded: Other neonatal morbidity.*

**Maternal morbidity definition 1:** Any occurrence of assisted delivery, caesarean section, preeclampsia and eclampsia, placental disorders, or haemorrhage. *Excluded: Other maternal morbidity.*

**Neonatal morbidity definition 2:** Any occurrence of preterm birth, small for gestational age, large for gestational age, low APGAR score, birth trauma, respiratory disorders, congenital malformation, or other neonatal morbidity. *Excluded: none.*

**Maternal morbidity definition 2:** Any occurrence of assisted delivery, caesarean section, preeclampsia and eclampsia, placental disorders, haemorrhage, or other maternal morbidity. *Excluded: none.*

**Interpretation:** G3 and G4 grandchildren have lower risk of neonatal morbidity according to both definitions (Definition 1: $OR_{G3} = 0.89$, $OR_{G4} = 0.96$; Definition 2: $OR_{G3} = 0.89$, $OR_{G4} = 0.95$). Mothers of G3 and G4 grandchildren also have lower risk of maternal morbidity according to definition 1. However, an advantage was only observed in mothers of G3 grandchildren with definition 2 (Definition 1: $OR_{G3} = 0.90$, $OR_{G4} = 0.95$; Definition 2: $OR_{G3} = 0.95$, $OR_{G4} = 0.99$). A general pattern emerges in both analyses suggesting that maternal factors may dominate paternal factors in the transmission of advantage in maternal morbidities. However, the evidence was more mixed in neonatal morbidities, with contrasting findings in G3 grandchildren and G4 grandchildren.

***Figure S2.6.*** Regression results for analyses of composite neonatal and maternal morbidity in G3 grandchildren and G4 great-grandchildren and their parents (definition 1)

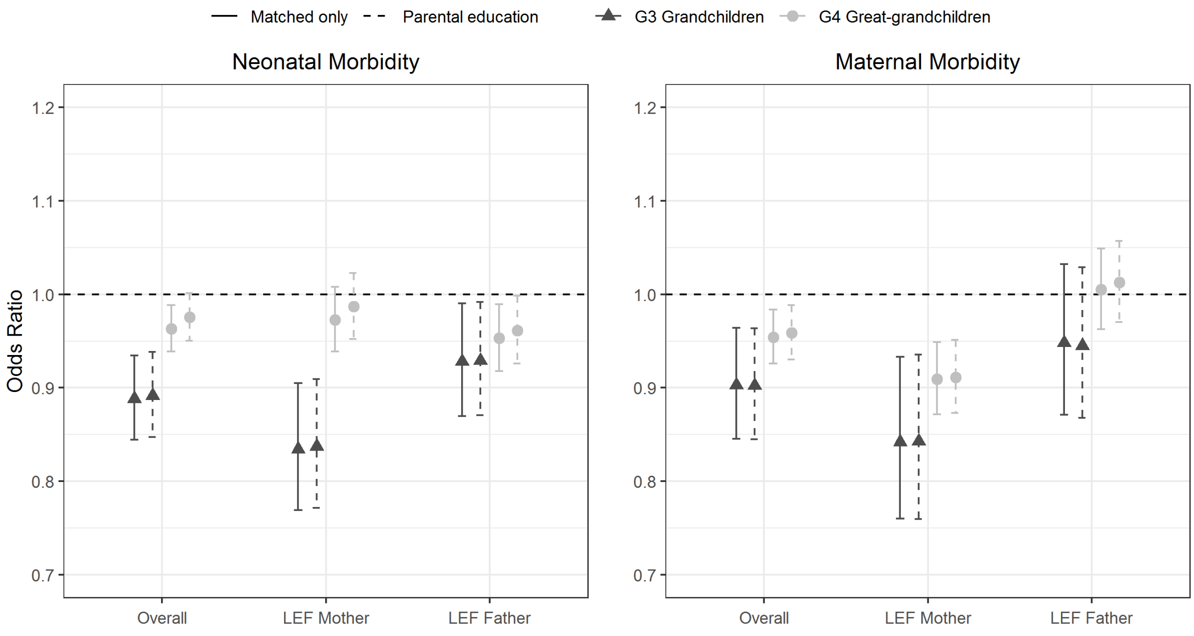


***Figure S2.7.*** Regression results for analyses of composite neonatal and maternal morbidity in G3 grandchildren and G4 great-grandchildren and their parents (definition 2)

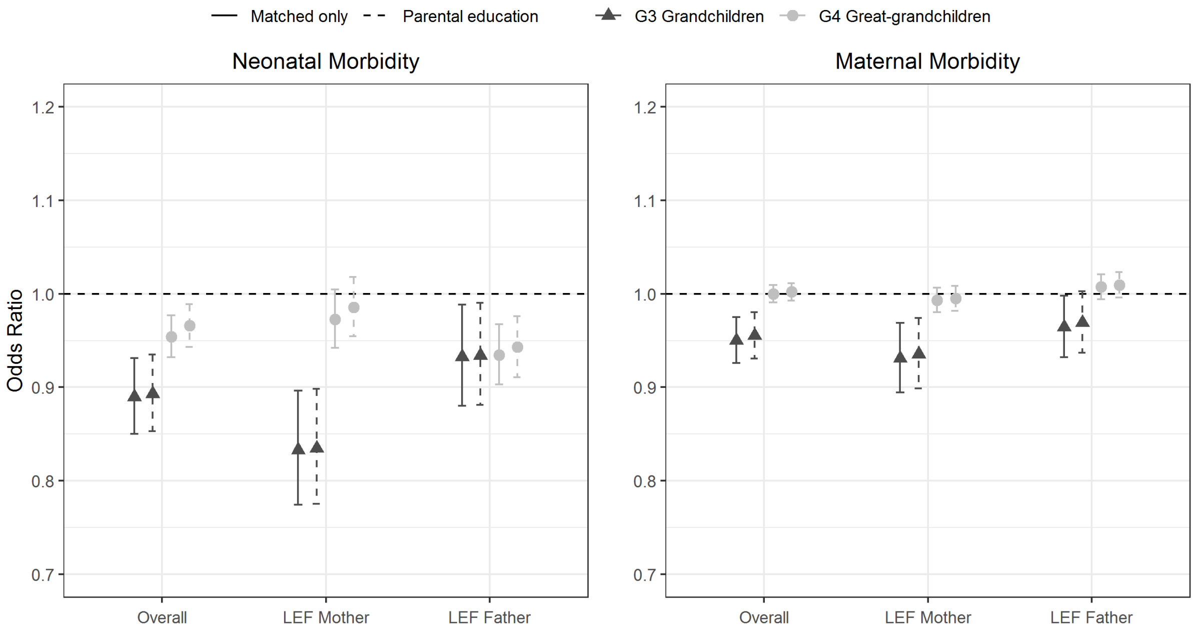

***Table S2.8.*** Regression results for analyses of composite neonatal and maternal morbidity in G3 grandchildren and G4 great-grandchildren and their parents (definition 1)

Neonatal morbidity - Definition 1

| Generation | Model | Adjustment | Odds Ratio | 95% CI | p |
|---|---|---|---|---|---|
| G3 | Overall | Matched only | 0.89 | [0.84, 0.93] | < 0.001 |
| | | Parental education | 0.89 | [0.85, 0.94] | < 0.001 |
| | LEF Mother | Matched only | 0.83 | [0.77, 0.91] | < 0.001 |
| | | Parental education | 0.84 | [0.77, 0.91] | < 0.001 |
| | LEF Father | Matched only | 0.93 | [0.87, 0.99] | 0.02 |
| | | Parental education | 0.93 | [0.87, 0.99] | 0.03 |
| G4 | Overall | Matched only | 0.96 | [0.94, 0.99] | 0.004 |
| | | Parental education | 0.98 | [0.95, 1] | 0.06 |
| | LEF Mother | Matched only | 0.97 | [0.94, 1.01] | 0.13 |
| | | Parental education | 0.99 | [0.95, 1.02] | 0.47 |
| | LEF Father | Matched only | 0.95 | [0.92, 0.99] | 0.01 |
| | | Parental education | 0.96 | [0.93, 1] | 0.04 |

Maternal morbidity - Definition 1

| Generation | Model | Adjustment | Odds Ratio | 95% CI | p |
|---|---|---|---|---|---|
| G3 | Overall | Matched only | 0.90 | [0.85, 0.96] | 0.002 |
| | | Parental education | 0.90 | [0.85, 0.96] | 0.002 |
| | LEF Mother | Matched only | 0.84 | [0.76, 0.93] | 0.001 |
| | | Parental education | 0.84 | [0.76, 0.94] | 0.001 |
| | LEF Father | Matched only | 0.95 | [0.87, 1.03] | 0.22 |
| | | Parental education | 0.95 | [0.87, 1.03] | 0.19 |
| G4 | Overall | Matched only | 0.95 | [0.93, 0.98] | 0.002 |
| | | Parental education | 0.96 | [0.93, 0.99] | 0.007 |
| | LEF Mother | Matched only | 0.91 | [0.87, 0.95] | < 0.001 |
| | | Parental education | 0.91 | [0.87, 0.95] | < 0.001 |
| | LEF Father | Matched only | 1.00 | [0.96, 1.05] | 0.82 |
| | | Parental education | 1.01 | [0.97, 1.06] | 0.57 |

***Table S2.9.*** Regression results for analyses of composite neonatal and maternal morbidity in G3 grandchildren and G4 great-grandchildren and their parents (definition 2)

Neonatal morbidity - Definition 2

| Generation | Model | Adjustment | Odds Ratio | 95% CI | p |
|---|---|---|---|---|---|
| G3 | Overall | Matched only | 0.89 | [0.85, 0.93] | < 0.001 |
| | | Parental education | 0.89 | [0.85, 0.93] | < 0.001 |
| | LEF Mother | Matched only | 0.83 | [0.77, 0.9] | < 0.001 |
| | | Parental education | 0.83 | [0.78, 0.9] | < 0.001 |
| | LEF Father | Matched only | 0.93 | [0.88, 0.99] | 0.02 |
| | | Parental education | 0.93 | [0.88, 0.99] | 0.02 |
| G4 | Overall | Matched only | 0.95 | [0.93, 0.98] | < 0.001 |
| | | Parental education | 0.97 | [0.94, 0.99] | 0.004 |
| | LEF Mother | Matched only | 0.97 | [0.94, 1] | 0.09 |
| | | Parental education | 0.99 | [0.95, 1.02] | 0.38 |
| | LEF Father | Matched only | 0.93 | [0.9, 0.97] | < 0.001 |
| | | Parental education | 0.94 | [0.91, 0.98] | < 0.001 |

Maternal morbidity - Definition 2

| Generation | Model | Adjustment | Odds Ratio | 95% CI | p |
|---|---|---|---|---|---|
| G3 | Overall | Matched only | 0.95 | [0.93, 0.98] | < 0.001 |
| | | Parental education | 0.96 | [0.93, 0.98] | < 0.001 |
| | LEF Mother | Matched only | 0.93 | [0.89, 0.97] | < 0.001 |
| | | Parental education | 0.94 | [0.9, 0.97] | 0.001 |
| | LEF Father | Matched only | 0.96 | [0.93, 1] | 0.04 |
| | | Parental education | 0.97 | [0.94, 1] | 0.07 |
| G4 | Overall | Matched only | 1.00 | [0.99, 1.01] | 0.99 |
| | | Parental education | 1.00 | [0.99, 1.01] | 0.67 |
| | LEF Mother | Matched only | 0.99 | [0.98, 1.01] | 0.31 |
| | | Parental education | 1.00 | [0.98, 1.01] | 0.46 |
| | LEF Father | Matched only | 1.01 | [0.99, 1.02] | 0.28 |
| | | Parental education | 1.01 | [1, 1.02] | 0.16 |

#### 3. Sensitivity and Robustness Analyses

Items S3.1 and S3.2 describe sensitivity analyses we undertook to assess the robustness of our 'overlapping generations' analyses, where we compared G3 grandchildren to G4 great-grandchildren directly within their overlapping birth cohort periods. The analyses below introduce controls into the mix and use them in specific analyses as negative controls. In all these analyses, the full matching criteria used as adjustment covariates. Below we provide the rationale for each analysis and our interpretation:

**LEF-G3 vs LEF G4:** This analysis is what was presented in the main paper.

**LEF-G3 vs Control-G4:** Since we see no difference in infant mortality between G4 great-grandchildren and their controls (See **Figure 3**), we hypothesised that comparing G3 grandchildren to the matched controls of the G4 great-grandchildren in the same overlapping period analysis, we would observe the same estimate as when comparing to the G4 great-grandchildren directly. This hypothesis was confirmed (LEF-G3 vs LEF G4 HR = 0.32, LEF-G3 vs Control-G4 HR = 0.33).

**LEF-G4 vs Control-G3:** Similarly, since G4 great-grandchildren exhibit no advantage compared to the general population, we would expect to observe no advantage when comparing them in the overlapping generation analysis to the matched controls of G3 grandchildren. This hypothesis was confirmed (LEF-G4 vs Control G3 HR = 0.99).

**Control-G3 vs Control-G4:** The above analyses establish that the selected controls behave in such a way to imply that direct comparisons between G3 grandchildren and G4 great-grandchildren would be unbiased. Similarly, the findings also imply that if we compared matched controls of G3 grandchildren to matched controls of G4 great-grandchildren, we would observe no mortality differences. This hypothesis was again validated (Control-G3 vs Control-G4 HR = 1.21). Note that there was some model instability when adjusting for parental education level in this specific analysis, evidenced by dramatically narrower confidence intervals. This is unusual behaviour when including additional covariates which do not meaningfully change the point estimate. We were unable to identify an exact cause of this problem.

If the trend for increased mortality in G3 controls vs G4 controls 3 is to be interpreted literally despite large confidence intervals, this would imply that our analyses are biased towards generations having a survival advantage. This in turn would imply that our estimates showing a survival advantage in G3 compared to G4 are in fact conservative. However, we do not think this is likely, given the results of the other negative control analyses. Nonetheless, this possibility does conserve our interpretation of a strong dilution of the survival advantage between generations G3 and G4 in longevity-enriched families.

Overall, these findings suggest that our direct comparison analyses in the main article are capable of accurately inferring patterns in intergenerational transmission of infant survival advantages, independent of both secular trends in the background population, and methodological and statistical considerations.

***Figure S3.1.*** Regression results for negative control analyses using overlapping generations and matched controls

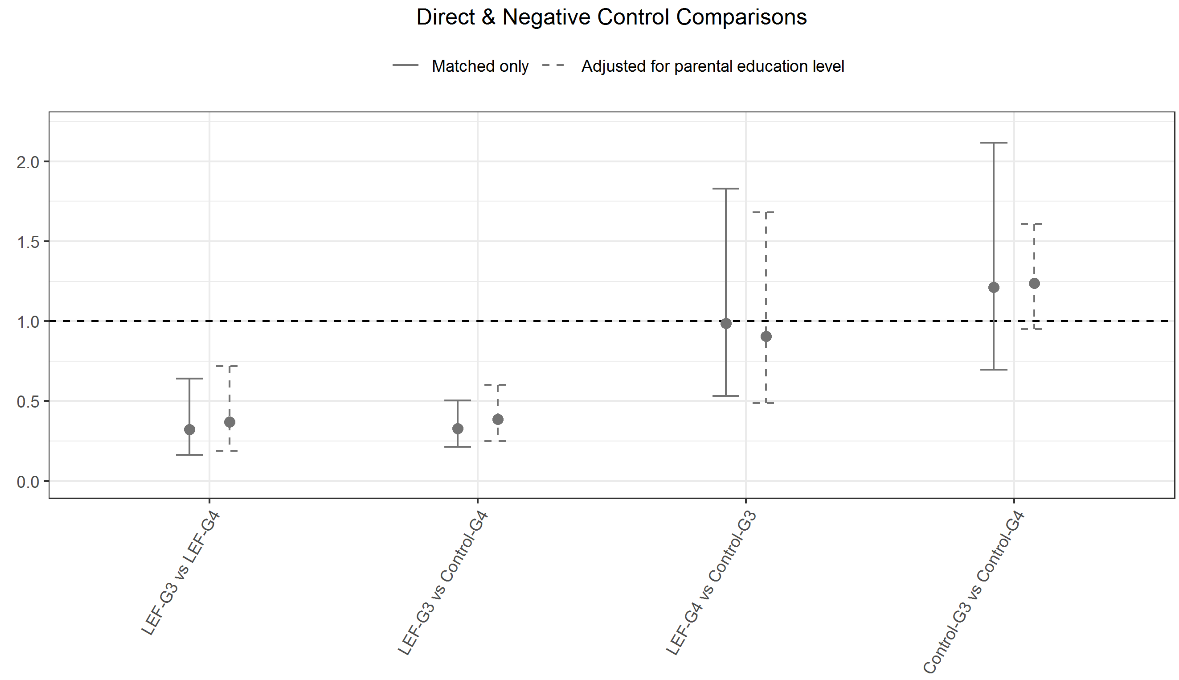


***Table S3.2.*** Regression results for negative control analyses using overlapping generations and matched controls

| Model | Adjustment | HR | 95% CI | p |
|---|---|---|---|---|
| LEF-G3 vs LEF-G4 | Matched only | 0.32 | [0.16, 0.64] | 0.001 |
| LEF-G3 vs LEF-G4 | Parental education | 0.37 | [0.19, 0.72] | 0.003 |
| LEF-G3 vs Control-G4 | Matched only | 0.33 | [0.21, 0.5] | < 0.001 |
| LEF-G3 vs Control-G4 | Parental education | 0.39 | [0.25, 0.6] | < 0.001 |
| LEF-G4 vs Control-G3 | Matched only | 0.99 | [0.53, 1.83] | 0.96 |
| LEF-G4 vs Control-G3 | Parental education | 0.91 | [0.49, 1.68] | 0.75 |
| Control-G3 vs Control-G4 | Matched only | 1.21 | [0.7, 2.12] | 0.5 |
| Control-G3 vs Control-G4 | Parental education | 1.24 | [0.95, 1.61] | 0.11 |

***Table S3*.3.** Regression results for analyses with varying windows of opportunity for the diagnosis of congenital malformations

| Generation | Time Window | Adjustment | Odds Ratio | CI | p |
|---|---|---|---|---|---|
| G3 | < 28 days | Matched only | 0.94 | [0.79, 1.12] | 0.49 |
| | | Parental education | 0.94 | [0.78, 1.12] | 0.46 |
| | < 60 days | Matched only | 0.98 | [0.84, 1.16] | 0.84 |
| | | Parental education | 0.98 | [0.83, 1.16] | 0.8 |
| | < 90 days | Matched only | 1.00 | [0.85, 1.17] | 0.98 |
| | | Parental education | 1.00 | [0.85, 1.17] | 0.98 |
| | < 365 days | Matched only | 0.98 | [0.85, 1.14] | 0.81 |
| | | Parental education | 0.99 | [0.85, 1.15] | 0.87 |
| | < 730 days | Matched only | 0.97 | [0.85, 1.12] | 0.7 |
| | | Parental education | 0.99 | [0.86, 1.14] | 0.85 |
| | > 730 days | Matched only | 0.97 | [0.87, 1.08] | 0.56 |
| | | Parental education | 0.98 | [0.88, 1.1] | 0.77 |
| G4 | < 28 days | Matched only | 0.92 | [0.84, 1.01] | 0.07 |
| | | Parental education | 0.94 | [0.86, 1.02] | 0.15 |
| | < 60 days | Matched only | 0.93 | [0.86, 1.01] | 0.1 |
| | | Parental education | 0.95 | [0.87, 1.03] | 0.2 |
| | < 90 days | Matched only | 0.93 | [0.86, 1.01] | 0.07 |
| | | Parental education | 0.94 | [0.87, 1.02] | 0.14 |
| | < 365 days | Matched only | 0.95 | [0.89, 1.01] | 0.12 |
| | | Parental education | 0.96 | [0.9, 1.03] | 0.23 |
| | < 730 days | Matched only | 0.97 | [0.91, 1.03] | 0.25 |
| | | Parental education | 0.98 | [0.92, 1.04] | 0.48 |
| | > 730 days | Matched only | 1.00 | [0.95, 1.05] | 0.91 |
| | | Parental education | 1.02 | [0.97, 1.07] | 0.55 |

**Interpretation:** no differences were observed across any of the diagnostic windows for assessing congenital malformations. Descendants of longevity-enriched sibships do not exhibit a lower risk of congenital malformations, at least in generations 3 and 4.

***Table S3.4.*** Regression results for analyses of mortality when adjusting for paternal immigration status and age at birth

| Generation | Adjustment | Hazard Ratio | CI | p |
|---|---|---|---|---|
| G3 | Matched only | 0.61 | [0.41, 0.91] | 0.015 |
| | Education | 0.65 | [0.43, 0.98] | 0.038 |
| | Education, Paternal Age | 0.65 | [0.43, 0.98] | 0.041 |
| | Education, Danish Father | 0.64 | [0.42, 0.97] | 0.036 |
| | Education, Paternal Age, Danish Father | 0.62 | [0.41, 0.93] | 0.022 |
| G4 | Matched only | 0.95 | [0.73, 1.24] | 0.713 |
| | Education | 0.99 | [0.75, 1.30] | 0.936 |
| | Education, Paternal Age | 0.99 | [0.76, 1.30] | 0.955 |
| | Education, Danish Father | 0.98 | [0.75, 1.28] | 0.884 |
| | Education, Paternal Age, Danish Father | 0.98 | [0.75, 1.29] | 0.899 |

**Interpretation:** Note that the matched only estimate (HR = 0.61) is slightly different to the baseline estimate provided in the main study (HR = 0.53). This is because the above analyses only include children whose father's place of birth and age at conception is known. Overall, we observe that there were no differences in any of the estimates after adjusting for paternal age, paternal place of birth, or both.

***Table S3.5.*** Regression results for analyses assessing other outcomes using weight at birth

| Generation | Outcome | Adjustment | Coefficient | CI | pval |
|---|---|---|---|---|---|
| G3 | SGA | Matched only | 0.83 | [0.76, 0.9] | < 0.001 |
| | | Parental education | 0.85 | [0.78, 0.93] | < 0.001 |
| | vSGA | Matched only | 0.80 | [0.7, 0.9] | < 0.001 |
| | | Parental education | 0.84 | [0.74, 0.95] | 0.004 |
| | eSGA | Matched only | 0.66 | [0.53, 0.83] | < 0.001 |
| | | Parental education | 0.71 | [0.57, 0.89] | 0.003 |
| | LBW | Matched only | 0.83 | [0.76, 0.9] | < 0.001 |
| | | Parental education | 0.85 | [0.78, 0.92] | < 0.001 |
| | vLBW | Matched only | 0.82 | [0.63, 1.06] | 0.14 |
| | | Parental education | 0.86 | [0.65, 1.14] | 0.29 |
| | eLBW | Matched only | 0.86 | [0.54, 1.36] | 0.52 |
| | | Parental education | 1.04 | [0.63, 1.69] | 0.89 |
| G4 | SGA | Matched only | 0.90 | [0.85, 0.94] | < 0.001 |
| | | Parental education | 0.93 | [0.88, 0.98] | 0.004 |
| | vSGA | Matched only | 0.87 | [0.81, 0.93] | < 0.001 |
| | | Parental education | 0.91 | [0.85, 0.97] | 0.006 |
| | eSGA | Matched only | 0.85 | [0.75, 0.96] | 0.01 |
| | | Parental education | 0.89 | [0.78, 1.01] | 0.07 |
| | LBW | Matched only | 0.90 | [0.83, 0.97] | 0.008 |
| | | Parental education | 0.93 | [0.86, 1.01] | 0.09 |
| | vLBW | Matched only | 0.82 | [0.63, 1.06] | 0.14 |
| | | Parental education | 1.16 | [0.96, 1.4] | 0.12 |
| | eLBW | Matched only | 1.03 | [0.76, 1.41] | 0.83 |
| | | Parental education | 1.03 | [0.75, 1.4] | 0.87 |

Abbreviations: SGA (small for gestational age), vSGA (very small for gestational age), eSGA (extremely small for gestational age) , LBW (low birth weight), vLBW (very low birth weight), eLBW (extremely low birth weight)

**SGA**: Less than -1.282 SDs below the expected intrauterine weight for gestational age (10$^{th}$ percentile)
**vSGA**: Less than -1.645 SDs below the expected intrauterine weight for gestational age (5$^{th}$ percentile)
**eSGA**: Less than -2.326 SDs below the expected intrauterine weight for gestational age (1$^{st}$ percentile)
**LBW**: Less than 2500g at birth
**vLBW**: Less than 1500g at birth
**eLBW**: Less than 1000g at birth

**Interpretation:** Degree of advantage increased in both grandchildren and great-grandchildren when decreasing the percentile for intrauterine weights. For example, G3 grandchildren were 34% less likely to be extremely small for gestational age compared to controls, compared to 17% less likely for just being small for gestational age.

***Table S3.6.*** Regression results for analyses of key neonatal indicators as continuous rather than dichotomized outcomes

| Generation | Outcome | Adjustment | Beta | CI | pval |
|---|---|---|---|---|---|
| G3 | APGAR Score | Matched only | 0.03 | [0, 0.06] | 0.05 |
| | | Parental education | 0.03 | [0, 0.05] | 0.08 |
| | Birth Weight | Matched only | 44.81 | [25.33, 64.29] | < 0.001 |
| | | Parental education | 37.43 | [17.93, 56.93] | < 0.001 |
| | Gestational Age | Matched only | 1.06 | [0.52, 1.61] | < 0.001 |
| | | Parental education | 0.98 | [0.44, 1.53] | < 0.001 |
| G4 | APGAR Score | Matched only | -0.01 | [-0.03, 0] | 0.14 |
| | | Parental education | -0.01 | [-0.03, 0] | 0.07 |
| | Birth Weight | Matched only | 32.33 | [20.51, 44.14] | < 0.001 |
| | | Parental education | 24.97 | [13.15, 36.78] | < 0.001 |
| | Gestational Age | Matched only | 0.50 | [0.22, 0.78] | < 0.001 |
| | | Parental education | 0.39 | [0.11, 0.67] | 0.006 |

**APGAR Score:** Measured on a scale of 0-10
**Birth Weight:** Measured in grams
**Gestational Age:** measured in weeks

**Interpretation:** All estimates are supportive of findings presented in the main article.

***Table S3.7.*** Regression analyses for analyses of mortality including weakly matched sets who were previously unable to be matched according to the full matching criteria

| Cohort | Generation | Adjustment | HR | CI | p |
|---|---|---|---|---|---|
| Main Cohort | G3 | Matched only | 0.53 | [0.36, 0.77] | < 0.001 |
| | | Parental education | 0.55 | [0.37, 0.82] | 0.003 |
| | G4 | Matched only | 0.90 | [0.7, 1.17] | 0.43 |
| | | Parental education | 0.95 | [0.73, 1.23] | 0.67 |
| Fully Matched | G3 | Matched only | 0.57 | [0.39, 0.82] | 0.002 |
| | | Parental education | 0.60 | [0.41, 0.88] | 0.008 |
| | G4 | Matched only | 0.89 | [0.69, 1.15] | 0.37 |
| | | Parental education | 0.93 | [0.72, 1.2] | 0.58 |

**Interpretation:** Of the 141 unable to matched initially, we were able to match 137 of them with just birth year, maternal birth year and sex. Four were unable to be matched at all even with this basic criterion. When appending our new matched sets to our main data and reperforming the survival analyses, only minor changes in our estimates were observed (0.53 -> 0.57 in G3 grandchildren, 0.90 -> 0.89 in G4 great-grandchildren). We conclude that the unmatched persons excluded from our main cohort were missing at random and does not affect our inferences.

***Table S3.8.*** Missingness of data in key outcomes derived from the Medical Birth Registry stratified by levels of censoring variables

| Outcome | Infant Mortality | Not Missing | Missing | % Missing |
|---|---|---|---|---|
| Birth Weight | Alive | 60743 | 642 | 1.05 |
| | Dead | 302 | 16 | 5.03 |
| Gestational Age | Alive | 54311 | 7074 | 11.52 |
| | Dead | 256 | 62 | 19.50 |
| APGAR Score | Alive | 55046 | 6339 | 10.33 |
| | Dead | 241 | 77 | 24.21 |
| Maternal Smoking | Alive | 39696 | 21689 | 35.33 |
| | Dead | 152 | 166 | 52.20 |

**Interpretation:** Several of the outcomes used in our study which were derived from the Medical Birth Registry had patterns of missingness that were not random. For birth weight, gestational age, APGAR score, and maternal smoking, the rate of missingness was higher in children who died in the first year of life. Since children descending from longevity-enriched sibships have a lower rate of infant mortality (HR = 0.53), this implies our analyses assessing differences in adverse outcomes such as low birth weight, low gestational age, low APGAR score, and maternal smoking, are affected by a bias that goes in the direction of the observed advantages in these outcomes being conservative.
